# Simulating a potential mpox outbreak: Implications for control in non-endemic settings

**DOI:** 10.1101/2025.06.29.25330499

**Authors:** Philip Cherian, Gautam I. Menon

## Abstract

We construct a model for mpox outbreaks in low and middle-income countries where the disease is non-endemic, using India as an example. We simulate potential outbreak scenarios using BharatSim, a flexible agent-based simulation framework. The spread of mpox is modelled as being driven primarily through sexual contacts within a subnetwork of men who have sex with men (MSM), embedded within a larger network representing their household and workplace contacts. Our model allows for disease spread through this expanded network, including the possibility of heterosexual transmission. We quantify the outbreak size, the dynamics of infection within and outside the MSM subnetwork, and the implications of vaccination and prophylactic measures for curtailing disease spread. These results should inform planning and policy measures for mpox control in generic LMIC settings.

**Author summary:** Mpox continues to be classified as a Public Health Emergency of International Concern. While the disease is very largely sexually transmitted, through networks of men who have sex with men (MSM), there is an emerging consensus that household and workplace transmission could expand the scope of a potential outbreak. Relatively little is known about mpox transmission in non-endemic settings, where disease reservoirs are absent. We construct and simulate an agent-based model of disease spread for a potential mpox outbreak in low and middle-income countries, using India as an example. Our model uses a network description of the population, with an embedded subnetwork of MSMs. Transmission events in both homes and workplaces lead to the spread of the disease on the extended network. Our results should inform potential policy interventions, including targeted vaccination programs, for mpox control.

## 1 Introduction

The mpox virus (MPV, MPXV, or hMPXV) is a zoonotic virus in the genus *Orthopoxvirus*, a genus that contains the variola, cowpox, and vaccinia viruses. First identified in humans in 1970, mpox is predominantly animal-borne, with a relatively small number of cases in humans and limited person-to-person transmission [1]. Since 2014, however, the annual number of human mpox cases in Africa has shown an increasing trend [1]. This secular shift has been ascribed to waning population immunity from the discontinuation of the smallpox vaccine, the emergence of new sub-clades, as well as changing environmental and social factors, including deforestation, armed conflicts, and migration [2].

Between 2018 and 2021, outbreaks in human populations were reported in six African countries, with the majority of cases in the Democratic Republic of Congo (DRC) and Nigeria. Until 2022, the spread of mpox was largely limited to Africa, with only 58 reported cases being detected outside the continent. Of these, the largest outbreak (47 cases) was in the United States in 2003 [3]. The remaining cases were all connected to travel from Nigeria. No compelling evidence was found of person-to-person transfer of the disease, although two cases of secondary transmission were recorded [1]. Since 2022, disease spread has broadened to countries where mpox has not historically been endemic, with the current outbreak being the largest in history [4, 5]. The WHO declared mpox a Public Health Emergency of International Concern (PHEIC) in July, 2022. In the period between 01 January, 2022 to 31 May, 2025, 127 countries have reported a total of 150,092 mpox cases to the WHO. There have also been 343 confirmed deaths ascribed to mpox [6].

Historically, mpox infections in humans were thought to arise almost exclusively from contact with animal reservoirs. However, human-to-human transmission through direct routes (skin-to-skin contact, bodily fluids, and respiratory droplets) has also been recorded. Transmission could plausibly occur through exposure to skin lesions, droplets, and fomites [7, 8]. The 2022 outbreak of mpox was primarily associated with close intimate contact, including sexual activity. Most cases were diagnosed among men who have sex with men (MSMs), with MSMs constituting 98% of patients in a report of 528 cases from 16 countries [9, 10].

The mpox virus is associated with two primary clades, I and II, each divided into sub-clades a and b [11]. Clade I corresponds to what was earlier called the “Congo Basin clade”, while clades IIa and IIb correspond to the “West Africa clade”. Unlike clade Ia, which is primarily driven by exposure to animals and which has had limited potential for person-to-person transmission, clade Ib has also been reported to spread through sexual contact. Despite many uncertainties regarding the new strain, the observed shift in the modes of transmission, coupled with its rapid spread, prompted the WHO to redeclare mpox as a PHEIC in August, 2024 [12].

A very recent outbreak in Sierra Leone is the largest ever recorded in the country and the first major epidemic caused by the clade IIb mpox virus in Africa [13–15]. Interestingly, females and males have been equally affected in Sierra Leone (47·5% female and 52·5% male), a pattern that contrasts with data from historical clade IIb outbreaks in west Africa [13]. This may indicate alternate routes for transmission beyond the MSM route. The first mpox case in India, a country where mpox is not endemic, was recorded on 14th July, 2022, from an individual who had returned from the United Arab Emirates. Since then, over 30 mpox cases have been reported [16], with one recorded fatality, a 22-year-old male who had been in contact with a confirmed case of mpox in the UAE [17]. Almost all cases of mpox in India have been restricted to Clade IIb. However, in September 2024, India recorded its first reported case of the more virulent Clade Ib [18].

Guided by concerns over the possibility of enhanced human-to-human spread of mpox and the need to have appropriate modelling frameworks in place in advance of a potential outbreak, we report a modelling study of an mpox outbreak both within and outside MSM sexual networks. We concentrate on settings where mpox is non-endemic, thereby excluding the possibility of environmental reservoirs that might sustain endemicity. We simulate potential scenarios of such an mpox outbreak using BharatSim, an agent-based simulation framework constructed for the Indian population [19].

Although BharatSim was developed to study disease spread in India, the results presented here should be largely applicable to other LMICs. We note that while statistical models that address related questions have been formulated and studied [7], our work provides the first detailed agent-based model description of a potential mpox outbreak in India and, by extension, in other LMICs.

Our model population contains both MSMs and the network of their contacts. Drawing on published data describing sexual contact networks, we create a synthetic population of agents that represents a community of MSMs and their household and workplace contacts. If transmission does not occur between the MSMs and non-MSM populations, the spread of the disease is confined to the sexual contact network alone. We explore the potentially more serious consequences, at the population level, of weak transmission between an infected member of an MSM subnetwork and their non-MSM household and workplace contacts, from where the disease can spread further. Disease transmission occurs in the background of mobility between households and workplaces, of sexual encounters within the MSM subnetwork, as well as the possibility of transmission through heterosexual contacts.

We address three questions: (i) How does the network structure of sexual and non-sexual physical contacts in a typical non-endemic LMIC setting determine the epidemic dynamics of an mpox outbreak, (ii) How does the disease expand to non-MSM networks, given contacts at homes and workplaces, and (iii) What strategies might be most effective in curtailing the spread of the disease?

## 2 Methods

### 2.1 Model

The individuals in our model are part of a network, and move between different locations during a single day. Individual agents spend 12 hours at their homes, interacting with household contacts, and then move to 12 hours at their workplaces or schools to interact with workplace and school contacts. Individuals above the age of 18 are assumed to be employees who work at a workplace, while the remaining are assumed to be children who go to schools. At any given time, each location is assumed to be well-mixed, with all agents in that location in contact with all other agents in that location at that time. The model and associated population files have been made available on the GitHub repository https://github.com/dpcherian/mpox-model.

### 2.2 The disease progression

Following existing literature [20–22], we model mpox using compartments of an *SEIR* model, with Susceptible, Exposed, Infected, and Recovered individuals, as shown in Fig. 1. The latent period for the disease is assumed to be 7 days, and the infectious period to be 21 days (following, for example, Refs. [1, 7, 23–25]). Sojourn times in each of these compartments are assumed to follow an exponential distribution. We ignore the possibility of reinfections. (The existing literature does not seem to provide us a consistent picture regarding reinfections, with the distinction between a relapse and a reinfection being somewhat blurred [26]). We allow for a tunable fraction of breakthrough infections following a one-shot vaccination, choosing a figure of 75% efficacy for concreteness. This number has been chosen to correspond to the efficacy of a single-shot JYNNEOS vaccine [27–29], widely authorised and deployed during recent mpox outbreaks [30, 31].

**Fig 1.**
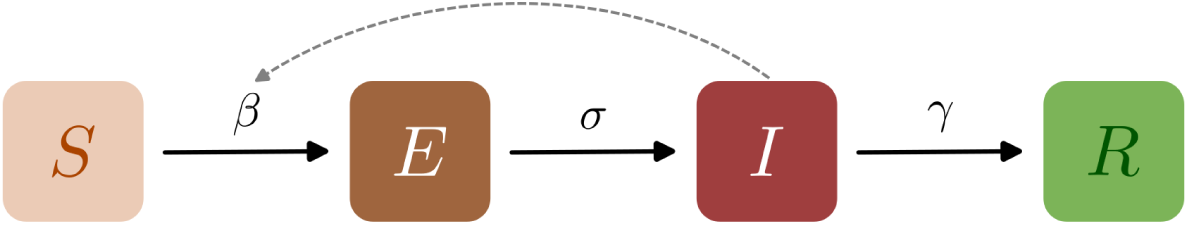
Schematic of the epidemiological model. Boxes show the set of states that characterise each individual in the population. Each individual can be in one of four compartments: (S)usceptible, (E)xposed, (I)nfected, and (R)ecovered. Black arrows indicate possible transitions between these states, while the dashed line indicates that the rate of infection of susceptible individuals is an increasing function of the number of infected individuals they are in contact with. Recovered individuals are assumed to be immune to further infection in this model.

### 2.3 Description of the network structure

In addition to individuals moving between homes and workplaces in 12 hour intervals, MSM agents in our simulations can also meet other agents in their sexual contact network through a sexual “encounter”. The probability distribution of the waiting-time between sexual encounters for an individual MSM is assumed to follow an exponential distribution with a rate *λ*_sex_ = 1/7 day*^−^*^1^. This corresponds to an MSM initiating a sexual encounter on average once every *τ* = 7 days. Once an encounter is initiated, a random contact from their list of sexual partners is chosen.

During such a sexual encounter, if either one of the two MSMs is infected with the other being susceptible, the susceptible one can become infected, with some probability *µ*. In the absence of empirical data on this parameter, we vary *µ* from 0–100% in our simulations, as done in Ref [7]. Other modelling studies (see Ref [25], for example) assume a single value of 20%. We note that *µ* can also be used to model the usage of prophylactic measures such as condom use, which have been found to reduce the risk of mpox spread [32]. The parameter *µ* can be further calibrated if the secondary attack rate (SAR) of the disease is known.

Since granular data for sexual contacts for MSMs is hard to obtain – it could potentially place individuals at risk – our model reconstructs synthetic sexual networks described statistically using the aggregate data in Refs. [33] and [34]. Studies estimating the size of the MSM population in India generally suggest that roughly 0.1–0.3% of the population identify as MSM [35–37], numbers that are similar to those provided by the Government of India (through the Ministry of Health and Family Welfare and the National AIDS Control Organisation) in 2012 [38, 39]. We cannot discount the fact that these numbers are underestimates [40, 41].

Thus, of the 100,000 individuals in our population, 1% are assumed to be MSM, while the others are their household and workplace contacts. The households and workplaces are distributed so that their mean occupancies are 4 and 50 respectively. This is done by first creating 25,000 homes (corresponding to an average household size of 4 individuals) and assigning each individual to one of them with a uniform probability. The same process is repeated with workplaces, ensuring the correct average number of agents per location, distributed as a Poisson distribution.

### 2.4 Construction of the MSM sexual contact network

A quantity of some importance is the network of MSMs in the population. Studies have shown [7] that the sexual-networks of MSMs are often heavy-tailed, and are reasonably well modelled by distributions like the truncated power-law distribution. We use the range of number of partners shown in Ref [33], modelling this through a truncated power-law distribution, where the lowest and highest values in this range correspond to the values in Ref. [33]. The power-law exponent is chosen such that the median of the distribution is the same as that provided in Ref. [33].

In order to construct a sub-population of men who have sex with men (MSMs) within our population, we begin by choosing 1000 men above the age of 18 (1% of the total population) at random from the 100,000 agents. We designate these agents as MSMs. Each MSM in this population is linked to a set of other MSMs who represent their sexual contacts. If we consider each MSM to be a node in a graph, and the links between MSMs who are sexual contacts to be edges, this produces an undirected graph representing the network. We decide the number of these sexual contacts for each MSM agent by drawing a random number from a truncated power law distribution in such a way that it reproduces the aggregate data from Ref. [33]. This represents the degree distribution of the graph.

We choose a truncated power-law distribution because empirical studies of MSM sexual-contact networks consistently report substantial heterogeneity in the number of sexual partners, with most individuals having relatively few partners and a small fraction having disproportionately many [7, 42]. A power-law form captures this heavy-tailed structure, which is important for representing heterogeneous transmission and potential superspreading dynamics. Additionally, we impose lower and upper cut-offs to reflect the finite range of partner numbers observed empirically in Ref. [33]. The lower and upper cut-offs therefore represent the observed range of available data.

Since we have no information about the true degree distribution of sexual contact networks in the Indian context, we make certain assumptions. First, we assume the distribution of sexual contact networks to follow a truncated power law, of the form *P* (*k*) ∼ 1*/k^α^* for *k* ∈ (*k*_min_*, k*_max_), where *P* (*k*) is the probability of an agent having *k* sexual partners, and whose minimum and maximum degree *k*_min_ and *k*_max_ we choose to be 3 and 19 respectively. The remaining free parameter in the distribution is the exponent *α*. We choose this exponent by requiring that the median of the truncated power law have a value of 6. This leads to a choice of *α* = 1.5. Similar values have been proposed for sexual contact networks in other countries [42–44]. The minimum, maximum, and median degrees are all chosen from the descriptive statistics of regular sexual networks of MSMs from three districts in India [33].

We sample this distribution for each MSM, obtaining a number of sexual contacts for that agent. We then assign links between these nodes by generating a random graph that follows this degree distribution. This is done using a sequential algorithm [45], implemented using the NetworkX package in Python [46]. This algorithm produces almost uniform random graphs with a prescribed degree sequence.

We have modelled the MSM network as a single subnetwork of the network defining the full population. In reality, multiple small disconnected components may be a better description. Our results thus explore a “worst-case” scenario, since the entire MSM subnetwork is susceptible to infection that begins at any node within it.

The resulting distribution of contacts for MSMs and their graph is shown in Fig. 2. This should be compared to similar figures in Ref. [33].

**Fig 2.**
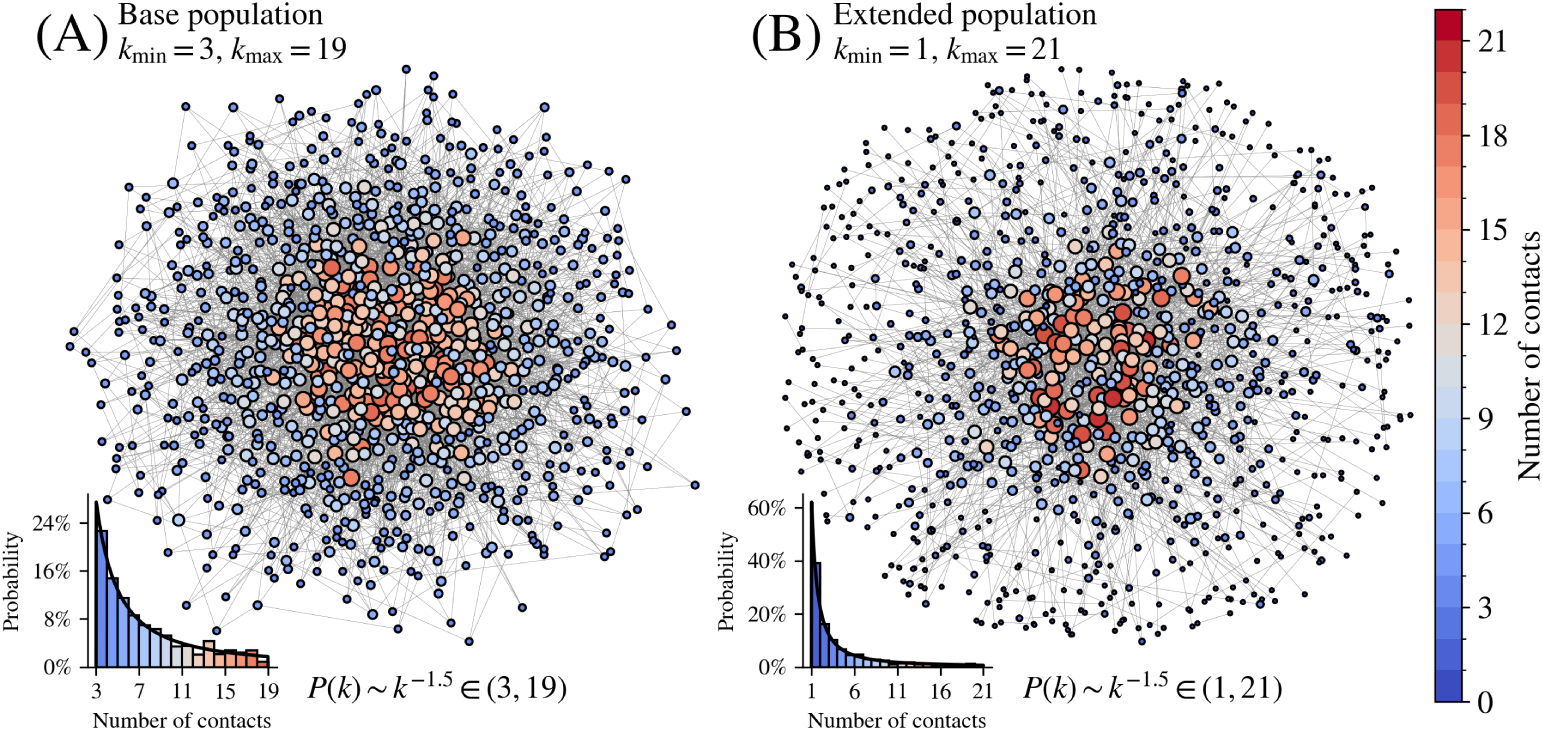
Graphs of two MSM subnetworks. Individual nodes with unique labels represent different MSM agents. (A) The population used in this paper. The degree distribution is assumed to be a truncated power-law distribution where the truncated values (3, 19) and the power-law exponent *α* = 1.5 were chosen to agree with Ref. [33]. In the inset we show the distribution of contacts, compared with the theoretical distribution. (B) An extended network using the same power-law exponent but truncated to (*k*_min_*, k*_max_) ≡ (1, 21). The extended network has many more low-degree nodes, resulting in a network with a high-degree “core”. The results from this extended population are shown in Appendix S1.

The network shown here has no nodes of degree less than 3, consistent with the data provided in Ref. [33]. We attribute this to their sampling strategy, in which each node (initial participant) was required to bring in three sexual partners separately as participants, a purposive sampling method called respondent-driven sampling. By construction, nodes with fewer than three partners will then be excluded.

To address this, we have created networks where the data is modelled by the same power law as applied to the truncated data but extending it to both the initial excluded range (1–3) as well as the tail (19–21). Results for this modified network are provided in Appendix S1.

### 2.5 Disease transmission

Agents can transmit the disease to each other in one of two channels: either sexually (for MSMs) or non-sexually (for everyone). In the results that follow, sexual transmission between infected MSMs and generic non-MSM individuals, or between infected non-MSM individuals, is not explicitly modelled. Epidemiological surveillance data seems to indicate that most transmission chains reported are linked primarily to MSM networks [47]. Furthermore, sexual behaviours traditionally associated with higher risk of sexually transmitted infections, particularly condomless anal sex, were strongly associated with mpox infection risk. A seroprevalence study among MSM in Berlin found that the number of condomless anal sex partners was strongly associated with both diagnosed and undiagnosed mpox infection [48]. A systematic review and meta-analysis similarly reported that unprotected anal and oral sex was significantly associated with mpox risk [49]. Nevertheless, studies have shown that close to a third of the MSM population in India is bisexual [50, 51]. Of these, nearly 75% have a predominant female partner who is their wife. Consequently, we have also simulated a case in which 33% of MSMs are taken to be bisexual. For these bisexual MSM agents, in addition to their MSM contacts drawn from the distribution as described earlier, a single female household member is further designated as their female sexual partner. The results of these simulations are shown in Appendix S2.

We assume that the greatest transmission of the disease between agents via non-sexual contacts occurs at home. The parameter *β* controls the force of infection at home. This parameter is varied, to see how the spread of the disease varies with the disease transmission strength. In addition, the model also allows for a weak probability of transmission in the workplace as well, assuming a reduced level of physical contact leading to infection. We average over a large number of stochastic runs, indicated in the figure captions to each figure that reports simulation results.

**Table 1.** Parameter choices for our mpox model. The simulations in this paper were run for the range of parameter choices given above.

| Par. | Description | Value | Comment |
| --- | --- | --- | --- |
| $N$ | Total population size | 100,000 | MSM agents: 1000, non-MSM agents: 99,000. |
| $\beta$ | Household transmission parameter | (0.1, 0.3, 0.5) | Model choice. |
| $\sigma$ | Transition rate from exposed to infectious | $1/7 \text{ day}^{-1}$ | Ref. [1, 25]. |
| $\gamma$ | Transition rate from infectious to recovered | $1/21 \text{ day}^{-1}$ | Ref. [1, 23, 24]. |
| $\mu$ | Probability of infection following a sexual encounter | (0.0, 0.2, 0.4, 0.6, 0.8, 1.0) | Model choice. |
| $\lambda_{\text{sex}}$ | Rate at which MSMs initiate sexual contact | $1/7 \text{ day}^{-1}$ | Model choice. Sexual encounters are initiated on an average once a week. |
| VPD | Number of vaccines available per day | 0, 10, 100 | Corresponding to 0%, 1%, and 10% of the MSM population per day. |
| VE | Vaccine efficacy: reduction in probability of infection | 75% | From JYNNEOS vaccine efficacy [27–29]. |
| $\tau_{\text{SR}}$ | Average time to self-report symptoms | (0 days, 1 week, 2 weeks) | Model choice, informed by Refs. [52–54]. |
| $f_{\text{trace}}$ | Fraction of MSM contacts traced | (0.2, 0.4, 0.6, 0.8) | Model choice, informed by Refs. [55, 56]. |
| $\beta_{\text{work}}$ | Workplace transmission parameter | (0%, 1%, 5%) of each $\beta$ | Agents experience a transmission parameter of $\beta$ at homes, and $\beta_{\text{work}}$ at their workplaces. |

### 2.6 Introducing vaccination

We consider a single dose vaccine with an efficacy of 75%. We choose this value to reflect the effectiveness of a single dose of the JYNNEOS vaccine which was originally developed for smallpox but which has since been shown to protect against mpox as well [27–29]. Vaccination is assumed to have an immediate effect in reducing the relative risk of contracting the disease. Since little is currently known about the vaccine’s efficacy at preventing transmission or about the timescale over which vaccination becomes effective, we assume immediate protection and neglect any effect of vaccination in reducing the relative risk of transmission.

Vaccination in our simulations is restricted to the MSM subpopulation. This choice reflects epidemiological evidence from recent mpox outbreaks indicating that sustained transmission in non-endemic settings is primarily driven by dense sexual contact networks [7], within which MSMs constitute a core transmission group. In our model, transmission may occur outside this subnetwork through household and potentially through workplace contacts, but such transmission does not by itself sustain epidemic spread. Targeting vaccination to the MSM subnetwork therefore represents an efficient allocation of vaccines to the portion of the contact network that contributes most strongly to onward transmission. We emphasise that this assumption does not imply exclusive susceptibility of MSMs to mpox, but rather reflects a modelling choice aimed at evaluating targeted control strategies in settings where vaccine supply or operational capacity may be limited.

We compare three different vaccine strategies to infer how they influence the epidemic trajectory: random vaccinations, risk-based vaccinations, and ring vaccinations.

In the first strategy, MSMs are vaccinated at random with a fixed number of vaccines available daily. Thus, in this strategy, on any given day, all MSM agents who are not exhibiting symptoms are equally likely to be vaccinated. In the second “risk-based” strategy, MSMs are classified as “high” or “low” risk individuals, based on their number of sexual contacts in the MSM network. The lower 25% of the node degree distribution is assumed to be “low risk”, while the upper 25% are assumed to be “high-risk”. In Fig. 3, we show the degree distribution of contacts in our model population (the insets from Fig. 2), with this classification. Vaccination proceeds in descending order of risk class, with individuals selected at random within each class: high-risk agents are vaccinated first, followed by intermediate-risk and then low-risk agents.

**Fig 3.**
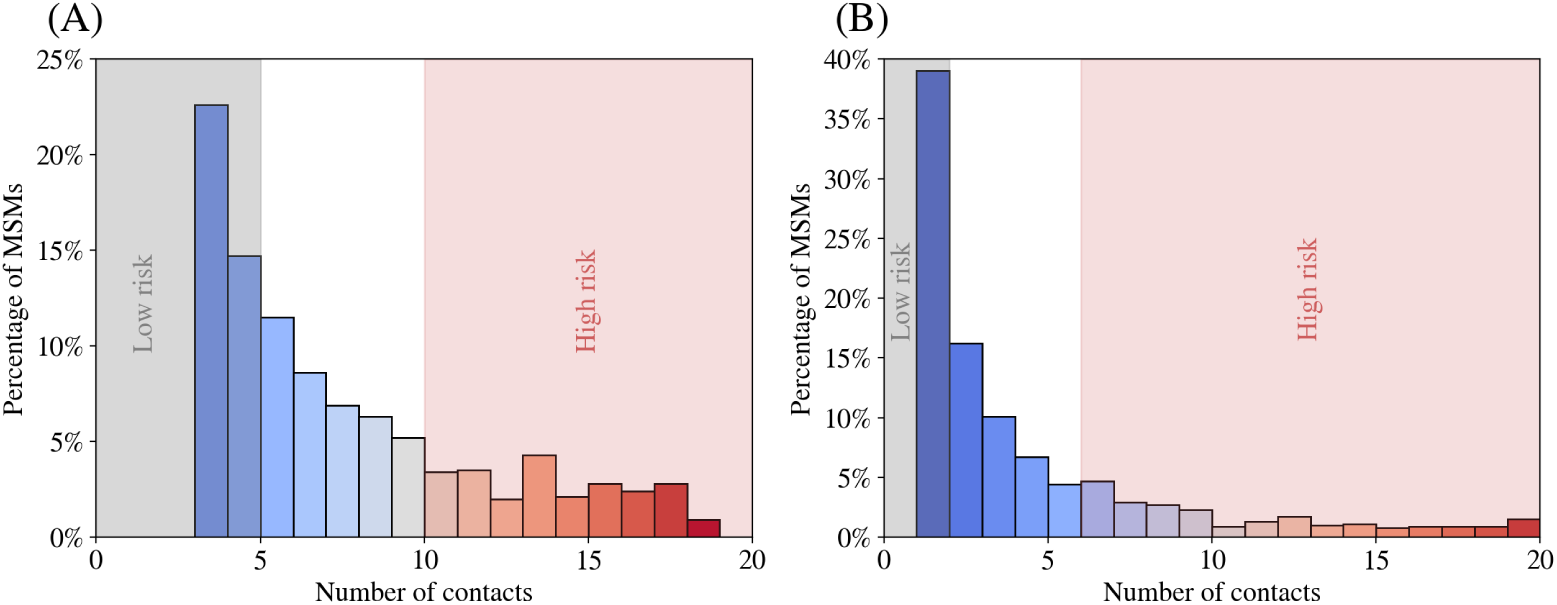
Classification of MSMs as low- and high-risk. Degree distribution of MSM agents in (A) the population used in the main paper, and (B) the extended population used for the simulations in Appendix S1. In each case, MSM agents are classified as low- or high-risk based on whether their number of sexual contacts lies in the lower or upper 25% respectively of all contacts. All other agents are classified as “intermediate” risk. The risk-based vaccination strategy first randomly vaccinates eligible agents from the high-risk class, followed by intermediate and then low-risk agents.

In addition to the random and risk-based vaccination strategies described above, we also consider a reactive “ring” vaccination strategy. In public health practice, ring vaccination involves identifying a set of individuals at elevated risk following the detection of an infectious case, and administering vaccines to their contacts and, where feasible, contacts of contacts [57]. This approach has historical roots in the global smallpox eradication campaign, where intensive contact tracing and vaccination of rings of exposure were used to interrupt transmission chains, supplementing broader immunisation efforts [58].

Ring vaccination has also been proposed and analysed in the context of mpox outbreaks [59–61]. In our simulations, ring vaccination is implemented as a reactive intervention triggered by the self-reporting of infected agents. We implement this strategy in our model as follows: When an MSM agent becomes infectious, he is assumed to self-report at some rate, which corresponds to a reporting delay *τ*_SR_ from the day that symptoms manifest. We simulate for three values of this average delay: immediate reporting, reporting after an average of 1 week, and reporting after an average of 2 weeks. Multiple studies of the 2022 mpox outbreak have shown that the mean reporting time of 1 week is reasonable, although some place this number slightly higher, up to 10–12 days [52–54]. We use the unrealistic “immediate” option as a counterfactual to simulate a “best-case” scenario.

Agents who recover before reporting never report their symptoms. Once an agent self-reports, a contact tracing process is initiated. Each reported individual has a subset of their recent contacts identified for vaccination, with the fraction of contacts successfully traced controlled by a contact tracing efficacy parameter *f*_trace_. All traced contacts are vaccinated immediately upon identification. This reflects a scenario in which sufficient vaccine doses are available to immunise all identified contacts, while allowing for imperfect tracing and delays in detection. As a result, the effectiveness of ring vaccination in the model is governed primarily by two operational factors: the speed with which infections are detected and the proportion of contacts that can be successfully traced. This is in contrast to the two earlier “pre-emptive” methods which had explicit constraints on vaccine supply.

## 3 Results

### 3.1 Unrestricted spread in MSM and non-MSM population

We simulate the spread of the disease by assuming an initial infection seed of a 10 MSM agents in the population. (Our simulations show that in order to get a comparable peak of infections starting with infections only in the non-MSM population, we would require a much larger number of initial infections; see Appendix S3. This has also been noted in earlier work [7].) In Fig. 4, we show the total infection curves for different values of the transmission parameter *β* and the probability of infection per sexual encounter *µ*. The different curves in each of the panels represent different values of *µ*. The figures show the mean over 500 simulations, but it is worth noting that there is substantial variance, with a good portion of runs dying out without percolating to the entire network.

**Fig 4.**
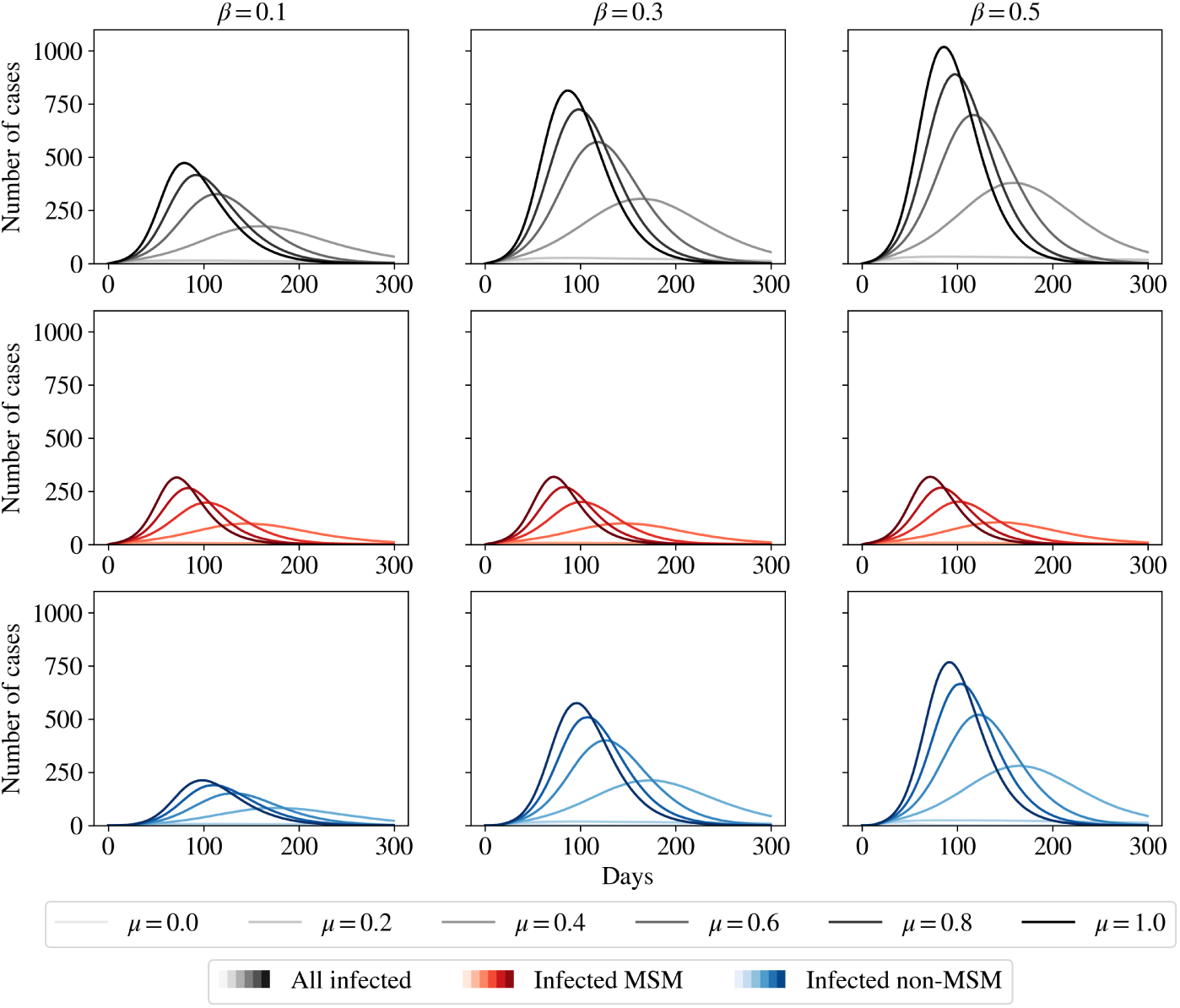
Active infections in the population. Infection curves for different values of household infectivity *β* and probability of sexual transmission *µ*, when the infection is seeded with 10 MSM agents. Each column shows the result for different values of *β*, while darker colours indicate higher values of *µ*. The top row of panels shows the total number of active infections in the population in each scenario. The second and third rows show the number of active infections in the MSM and non-MSM populations respectively, with the red curves showing the spread in the MSM population, and the blue curves showing the spread in the non-MSM population. The curves are averages over 500 runs.

In the top panel of Fig. 4, we show the epidemic curves for the total population for an initial infection seed of 10 MSM agents. We see that with increasing *µ* (the probability of transmission per sexual encounter), the epidemic peak is shifted earlier and is higher. The height of the peak also depends on the household transmission parameter *β*, which governs non-sexual transmission.

In the bottom panels of Fig. 4, we distinguish between the spread among the MSM and non-MSM population. We see that the peak in the general population is shifted further to the right as compared to the MSM peak. Additionally, we find that the MSM peak is more or less independent of the parameter *β*, when compared to the peak in the general population. However both of these peaks depend strongly on the parameter *µ*, the probability of transmission via the sexual channel.

Our results for the extended networks that have a larger number of lower degree nodes (shown in Appendix S1) are qualitatively identical. The peak is somewhat reduced, which leads to an overall lower outbreak size. We attribute this to the fact that single partner nodes are effectively protected from the disease if their partner remains uninfected or recovers before transmitting the disease. For higher degree nodes the number of ways in which disease can be contracted are higher and the likelihood of remaining uninfected correspondingly smaller.

When allowing for bisexual contacts, as shown in Appendix S2, we find that the spread in the MSM network is slowed down due to the existence of female sexual partners for a significant fraction of the MSM population. These female nodes for the bisexual MSMs act as “dead-ends” for the disease-spread, effectively slowing down the rate at which an infected MSM can infect their MSM sexual partners. The reduction in disease-spread in the MSM subnetwork leads to a smaller outbreak size among the non-MSM population as well. A notable exception occurs in the case of low *β* and high *µ*. For low *β*, the baseline household attack rate is relatively low in the absence of bisexual MSMs. Introducing bisexual individuals who may infect their female partners with a high *µ* effectively increases the number of household contacts exposed to infection, leading to a higher overall number of cases.

Lastly, in Appendix S4 we study the role that varying the frequency of sexual contact has on disease spread. We find that an increase in the meeting frequency can be offset by a reduction in *µ* to yield comparable overall transmission.

### 3.2 Effect of vaccination

In Fig. 5, we show the results for a vaccine drive with 10 vaccines per day for our population of 1000 MSMs. We show only the results for *µ* = 1.0; other values of *µ* show similar trends. We see that at this vaccination rate, vaccination has a large effect on reducing the peak of infections, and that risk-based vaccination is more effective than the random vaccination of MSMs. Vaccine administration both lowers the peak number of cases and shifts it to earlier times. However, as we show in Appendix S5, at higher vaccination rates (say, 100 vaccines per day), the difference between the risk-based and random vaccination strategies is negligible.

**Fig 5.**
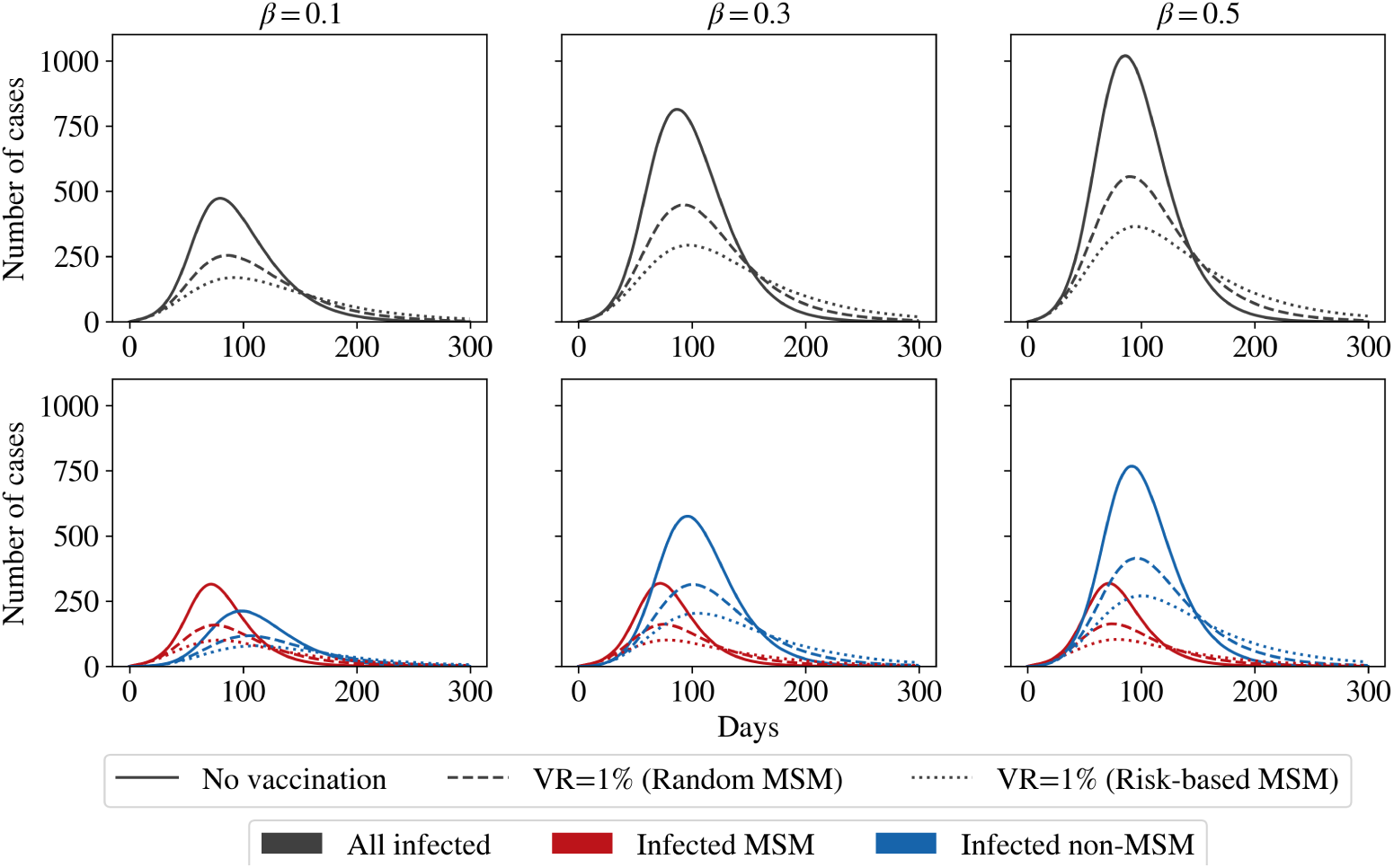
Effects of vaccination strategies on disease peak. A vaccination rate of 10 vaccines per day (VR=1% of MSMs) is very efficient in reducing the peak of infection, irrespective of strategy. However, the risk-based strategy of targeting “high-degree” MSMs is more efficient in bringing down the peak and shifting it earlier. Again, the curves are averages over 500 runs.

Despite the success of a risk-based vaccination strategy at lower vaccination rates, such a strategy may not be feasible, especially in the Indian context as it requires the explicit identification of “high-degree” individuals. In highly stigmatised epidemics like mpox among MSM populations, reliable information about sexual networks is difficult to obtain, and individuals with large numbers of contacts may be unwilling or unable to self-identify due to concerns about privacy, discrimination, or legal repercussions. Thus “pre-emptive” risk-based vaccination could face substantial operational and ethical barriers in such settings.

We can attempt to circumvent this using the ring vaccination strategy, where observed transmission events are used to implicitly target individuals at elevated risk. The effectiveness of ring vaccination emerges from the interaction of surveillance and contact tracing processes. Our results demonstrate that ring vaccination can reduce epidemic spread more than the random vaccination strategy, in specific parameter ranges. The benefits of ring vaccination are most pronounced under conditions of rapid case reporting and moderate contact tracing efficacy. In Appendix S6 we compare the effects of varying these parameters independently. We find that a moderate trade-off exists between the self-reporting rate and contact tracing efficiency: a higher reporting delay can be offset by a higher contact tracing efficiency, as shown in Fig. 6.

**Fig 6.**
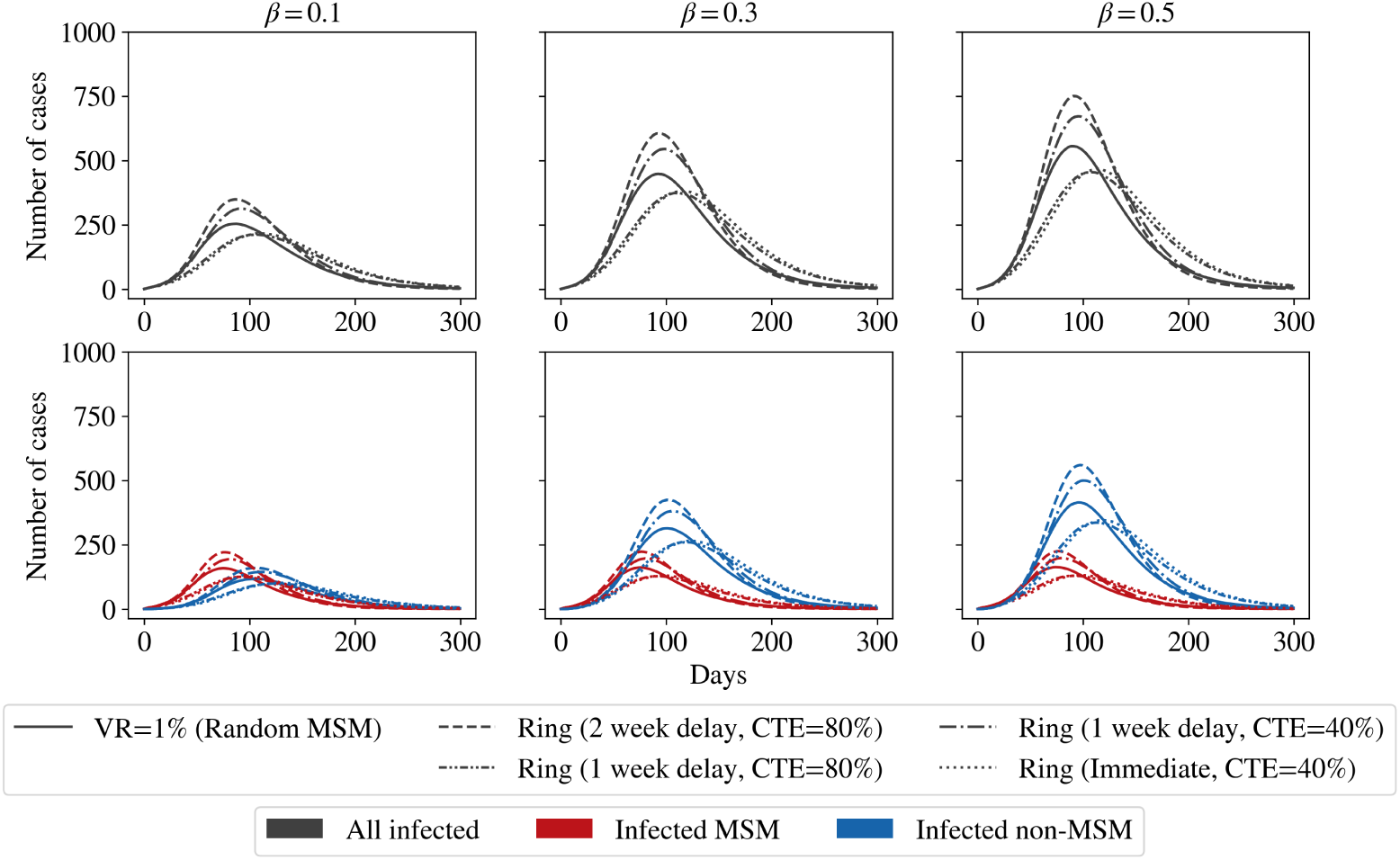
Trade-offs in ring vaccination. We compare ring vaccination strategies against random vaccination (with a vaccination rate of 1% of MSMs) for combinations of the mean self-reporting delay of *τ*_SR_, and contact tracing efficiency *f*_trace_. We find that in order for ring vaccination to be more efficient than random vaccination at these rates, the self-reporting delay must be low and the number of contacts traced must be high. However, the effect of a slower delay can be somewhat offset by an increased contact tracing efficiency. As before, the curves are averages over 500 runs.

At reasonable values of the reporting delay (1 week; see, for example, Ref. [52]) and contact tracing efficiencies of 40%-50%, ring vaccination can be found to have comparable results as random vaccination with a daily rate of 1% of MSMs per day. For shorter reporting delays and higher contact tracing efficacies, it can significantly outperform this random strategy. This strategy also has the effect of slowing down the speed of the epidemic, as the peak is shifted to later times.

Our findings align with real-world assessments of ring vaccination for mpox and related pathogens, which highlight the crucial role of timely case detection and rapid immunisation of contacts. For example, models of ring vaccination in the 2022 mpox outbreak show that reducing delays from symptom onset to diagnosis can dramatically lower the total number of infections, while higher contact coverage increases the proportion of cases averted [54]. At the same time, broad modelling of vaccination strategies suggests that combined strategies may outperform purely reactive ring vaccination alone when surveillance or tracing performance is limited.

### 3.3 Impact of workplace transmission

Finally, we explore the scenario of workplace transmission. These results are shown in Fig. 7, where the panels are the same as earlier figures, and the individual curves represent varying workplace transmission potential as fractions of *β*. Given reduced proximity at workplaces when compared to households, effective contacts at workplaces are expected to be much reduced compared to homes. We thus choose a reduced rate of infection (1% and 5% of the household rate *β*) in the workplace. Our results, in Fig. 7, show that the disease trajectory is significantly altered even at these low values of workplace transmission. When *β*_work_ is 5% of *β*, the peak is almost doubled, and a heavy infection tail is observed. The reasons for this are simply that while workplace infection is rare, once such an event occurs, the further possibility of household transmission leads to faster spread within the network.

**Fig 7.**
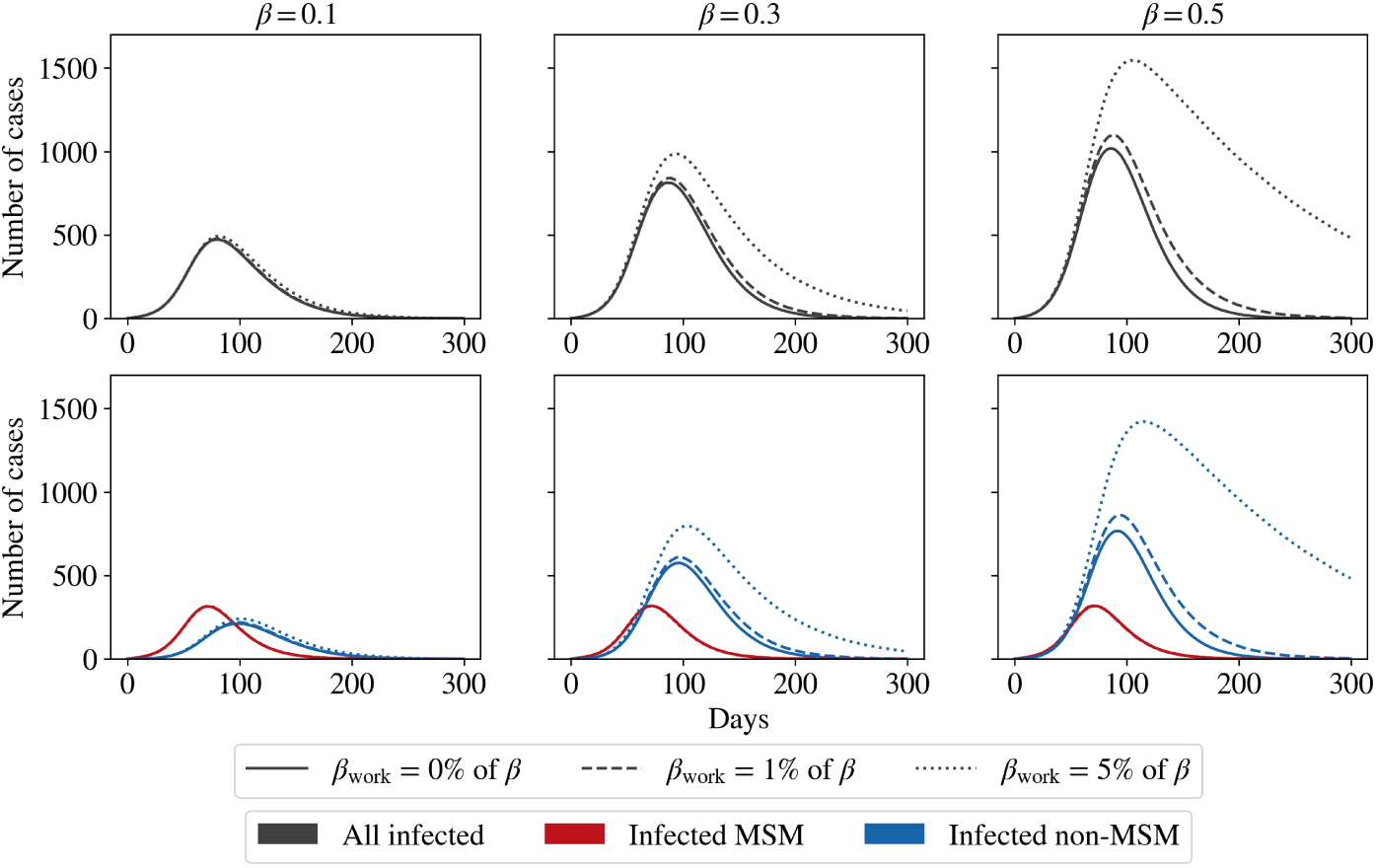
Effects of weak workplace transmission on active infections. While a workplace contact multiplier of 1% does not affect the peak significantly, a multiplier of 5% leads to a much higher peak and a much heavier tail. As before, the curves are averages over 500 runs.

Underlying these observations is the idea that the rapid spread in the smaller MSM network is what sustains the disease in the population. To verify this, in Appendix S7 we show the same curves as Fig. 7, but with *µ* = 0, meaning that we do not allow the disease to spread in the MSM subnetwork. We find that in this case, the disease dies out for all choices of parameters.

## 4 Discussion

We have described an agent-based model for the spread of mpox in overlapping networks of sexual and non-sexual interactions using BharatSim. For the parameters chosen here, consistent with estimated values from prior work, the disease spreads rapidly across the MSM subnetwork. The peak number of infections in the MSM population is directly sensitive to *µ*, the probability of infection per sexual encounter. Infections can expand beyond the MSM subnetwork at a small rate in our model, through contacts within the household and workplace.

As yet, the potential for sustained transmission of mpox within non-sexual networks is unclear. Preliminary data for the clade Ib was initially thought to reflect a substantial non-MSM component to infections. Current guidelines from multiple sources including the US CDC [62], the European CDC [63] and the World Health Organization [47] indicate that local person-to-person spread of clade I mpox has taken place in some non-endemic countries through day-to-day household contact, and within some healthcare settings, in addition to the normal spread of infections via direct sexual contact. Available evidence seems to suggest that such transmission outside non-MSM sexual networks is rare, though, and may not be a central driver of epidemic spread in non-endemic settings [47, 64], although occasional non-MSM sexual exposures have been reported. A survey of MSM with HIV/AIDS carried out in India showed that women whose sexual partners were MSM were at additional risk for HIV/AIDS [65]; such co-infections may also play a role in mpox spread but this is not modelled here.

The work described here models how infections can spread in non-endemic settings, showing how, depending on the efficiency of transmission in the MSM subnetwork and the broader contact network, policies for outbreak control may have to be applied to the larger contact network of MSMs. We found that restricting transmission to the household expands the numbers of infected. Adding the possibility of workplace transmission increases this number further, provided the rate of workplace transmission of infection is sufficiently high. Interestingly, these two modes of transmission, MSM and non-MSM, interact reciprocally to produce complex temporal behaviour [57]. While the disease cannot spread in purely non-MSM networks, since the relevant R_0_ is sub-threshold, the epidemic can be sustained through the interaction between networks.

The movement of the infection between MSM and non-MSM networks can lead to the emergence of two time scales in the progress of infection. We showed how disease spread in the population can be sustained even at very low infection seeds through spread in the MSM sexual network. A long tail of infection can be produced as a consequence of even weak spread involving workplace transmission. This is an interesting and, we believe, previously unanticipated result, which may have implications for the design of vaccination programs, in particular for how ring vaccinations might need to be structured. Our results also highlighted the importance of continued long-term surveillance, given these long tails in time.

We made specific approximations and assumptions regarding the nature of MSM networks in the Indian context, derived from a study in three Indian cities [33]. We used synthetic networks with the same statistics as those reported in these studies. One improvement on our model would be to incorporate real data on sexual contact networks, while protecting the privacy of individuals. We point out that a description similar to ours, of synthetic networks of MSM contacts has been used in Ref. [7]. Their networks, like ours, have heavy-tailed degree distributions, although the network properties differ in detail. The flexibility of our methods allows the networks our model describes to be changed easily.

Our networks are static and not dynamic, but some degree of dynamicity is to be expected, as members of the MSM population migrate in and out of the overall population, as well as establish new connections within pre-existing networks.

Agent-based models are powerful because they can be embedded into specific geographies, allowing for a description of physical hotspots of interaction, including casual sexual encounters. However, this would involve, at least for now, far more assumptions regarding the dynamical character of such MSM sexual networks – we defer these questions to future work. In this study, we modelled the latent period, infectious period, and self-reporting delay using exponential distributions, an assumption that simplifies the dynamics but does not fully capture the heterogeneity observed in clinical data. A natural extension of this work would be to incorporate empirically informed distributions for disease progression and reporting delays, which are often better described by lognormal distributions.

Work in Ref. [66] provides estimates of the numbers of the MSM population who arrange to meet via interactions on virtual platforms. To what extent this number constitutes a significant fraction of the total at-risk MSM population is unclear, although an important take-away from their study is that interventions that focus on physical hotspots will not reach the majority of MSM who use virtual sites. This observation is of relevance to potential intervention strategies. The Humsafar Trust, a non-governmental organization based in India, has reported an extensive study of the sexual and social networks of MSM and transgender/transsexuals (Hijras) across five Indian states [67]. Their work provides evidence for extensive sexual mixing and concurrent sexual relationships of MSM and Hijras, adding further layers of complexity that we ignore in this work. Their studies point to the role of community leaders, particularly in Hijra communities, in promoting safe sexual practices, a point of some relevance in deciding culturally and geographically appropriate interventions. Interventions in the case of a potential mpox outbreak should be sensitive to such social structures, for they may hold an important key to control.

Our work has specific relevance to mpox control in the Indian context, especially since the effectiveness of prophylactic measures such as vaccinations, applied to different sections of the population, can be easily assessed using BharatSim. We have, currently, no way of predicting which variant clade might transmit better through non-sexual routes, but believe that the expanding numbers of overall more transmissible variants highlights the need for adequate preparedness. Improved synthetic populations, better-calibrated MSM networks and a data-informed parametrisation of the overlap between sexual networks and other networks of physical contacts would enable a more refined description of how mpox might spread in the broader population. This would be essential to preparing for an mpox outbreak in the settings we consider. This is a possibility that cannot be discounted, given what we currently know.

## Data Availability

All data produced in the present work are contained in a freely accessible github repository at: github.com/dpcherian/mpox-model. The BharatSim code that produced these results is linked directly from the paper and is freely available for download.

## Acknowledgements

The authors are grateful for ongoing support from the Mphasis F1 Foundation. BharatSim development was initially supported by the Bill and Melinda Gates Foundation. GIM is a member of the National Disease Modelling Consortium, which is supported by the Gates Foundation (INV-044445). We are grateful to Drs. Tarun Bhatnagar, Venkatesan Chakrapani and Sheela Godbole for discussions. The authors acknowledge the use of computational facilities provided by the Centre for Bioinformatics and Computational Biology, supported by the DBT through grant no BT/PR40220/BTIS/137/22/2021, as well as the One-Health initiative of the Center for Climate Change and Sustainability at Ashoka University. The funders had no role in the study design, data collection and analysis, decision to publish, or preparation of the manuscript. Further, the results and conclusions presented here are based on research findings by the authors and are not the opinion of the Indian government.

## Supporting information

**Appendix S1 Simulations for extended sexual network.** We repeat the simulations from the main paper for a population with an extended MSM subnetwork generated from a truncated power-law degree distribution. We find that our qualitative results remain unchanged, but the values of the peak of the number of active infections are lower, since single-partner nodes are effectively protected if their partner remains uninfected or recovers before transmitting the disease. We also confirm that the effects of vaccination strategies and weak workplace transmission mirror those observed in the original population.

**Appendix S2 Effect of bisexual partnerships on disease spread.** We now study the effect of introducing female sexual partners for a subset of MSM agents in our population. Our results show that the inclusion of bisexual partnerships slows disease spread within the MSM subnetwork and consequently in the non-MSM network as well. In most parameter regimes, this also reduces the total number of non-MSM infections. However, when the household transmission parameter is low and the probability of transmission per sexual encounter is high, the increased household attack rate associated with infected female partners can instead lead to a modest rise in total non-MSM cases.

**Appendix S3 Simulations for an initial infection seed of non-MSM agents.** We consider the scenario in which the initial infection seed occurs in the non-MSM population, and find that in order to obtain a comparable disease peak, a substantially higher number of initial infections is required. These simulations exhibit a two-peak structure, with an initial non-MSM-driven outbreak followed by a second peak once transmission enters the MSM subnetwork.

**Appendix S4 Effect of sexual contact frequency on disease spread.** We study the effect of varying the frequency at which MSMs engage in sexual activity within their sexual network. We run simulations for a frequency of sexual contacts that is (i) double and (ii) half what is described in the main paper. We find, as expected, that increasing the frequency of sexual encounters effectively increases the rate of producing secondary infections. We also find that in the range considered here, the increase in frequency can be offset by a corresponding *decrease* in the probability of sexual transmission (i.e., *µ*) for the same reason.

**Appendix S5 Additional results for vaccination strategies.** We extend the vaccination analysis by considering higher daily vaccination rates with and without weak workplace transmission. We find that at sufficiently high vaccination rates the differences between strategies are effectively washed out, while lower rates substantially reduce, but do not eliminate, the infection peak.

**Appendix S6 Effects of self-reporting and contact tracing in ring vaccination.** We examine the impact of varying self-reporting delays and contact tracing efficiencies on the effectiveness of ring vaccination. We find that higher tracing efficiency significantly reduces and delays the infection peak, while shorter self-reporting delays improve outbreak control, although with diminishing returns at longer delays.

**Appendix S7 Primary and secondary transmission pathways.** We show results for the epidemic curves with workplace transmission without sexual transmission (*µ* = 0). We find that in all cases the disease dies out without spreading, demonstrating that the MSM subnetwork is essential for sustaining transmission in the population. Additionally, we classify active infections by infection source for the case where *µ* = 1.0. We find that the long tail in infections is driven by workplace contacts of MSMs and their households, which allows the disease to persist even after it has died out in the high-risk MSM subnetwork.

