## Supplemental Information for "Simulating a potential mpox outbreak: Implications for control in non-endemic settings"

### S1 Appendix: Simulations for extended sexual network

We consider the same population as the one described in the main paper, but connect the MSM agents using a different MSM sexual contact network. As described in the main paper, we generate connections between the MSMs with the same power-law exponent of $\alpha = 1.5$, truncated to the range $(1, 21)$. In this appendix we show the same results as in the main paper, but for this "extended" population, shown in Fig. 2B of the main paper.

In Fig. S1.1, we show the total number of active cases in the population, differentiating between cases in the MSM and non-MSM populations. The results are qualitatively identical to those described in the main paper, the only difference being that the number of cases is lower. This is a consequence of the fact that the number of infected MSMs is lower in this population, since the single-partner nodes are effectively protected from the disease if their partner remains uninfected or recovers before transmitting the disease.

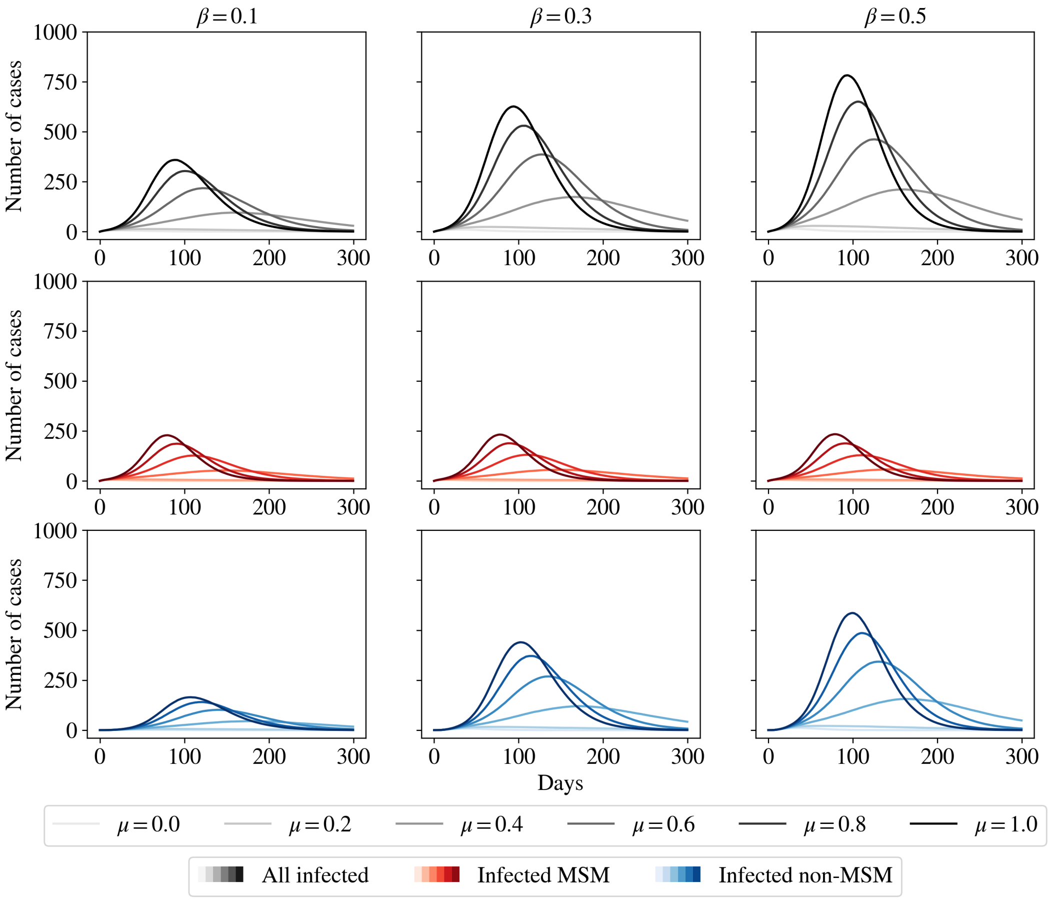


**Fig S1.1: Active infections in the population.** Infection curves for different values of household infectivity $\beta$ and probability of sexual transmission $\mu$, when the infection is seeded with 10 MSM agents, and the population is the extended population. Each column shows the results for different values of $\beta$, while darker colours indicate higher values of $\mu$. The panels show the total number of active infections, and the number of active infections in the MSM and non-MSM populations respectively. These results are qualitatively identical to those quoted in the main paper, but with significantly lower peaks. The curves are averages over 500 runs.

We also consider the case of different vaccination strategies for this extended population, with the same definition of "low" and "high" risk agents, as shown in Fig. 3B of the main paper. In Fig. S1.2, we show the effect of ten vaccines per day on the number of active cases. As before, the effect is qualitatively identical to that described in the main paper, with the exception of the peaks of all the curves which are much lower.

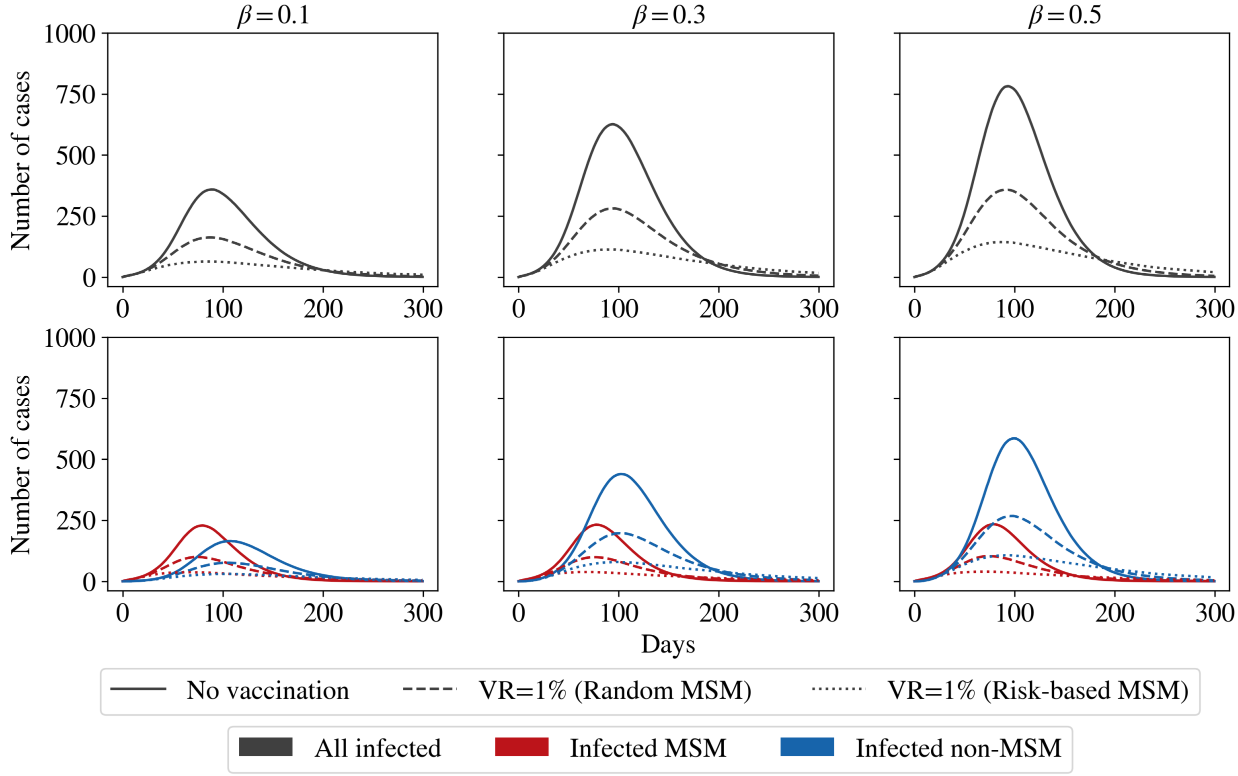


**Fig S1.2: Effects of vaccination strategies on disease peak for the extended population.** A vaccination rate of 10 vaccines per day (1% of MSM population) is very efficient in reducing the peak of infection, irrespective of strategy. However, the risk-based strategy of targeting "high-degree" MSMs is more efficient in bringing down the peak and shifting it earlier. These results mirror those from the original simulations. Again, the curves are averages over 500 runs.

Last, we consider the case of a small probability of workplace transmission, just as in the main paper. The results of these simulations, shown in Fig. S1.3, are again qualitatively identical to those described in the main paper: if $\beta_{\text{work}}$ has the relatively low value of 5% of $\beta$, a long tail of infection can be observed for higher values of $\beta$, although all peaks are lower than the corresponding values for the original population. In Fig. S1.4, we show the same curves while allowing for a weak workplace transmission but setting $\mu = 0$, meaning that no spread occurs in the sexual contact network. This result is identical to Fig. S7.1, as the only difference between the two populations occurs in the network structure of the MSMs.

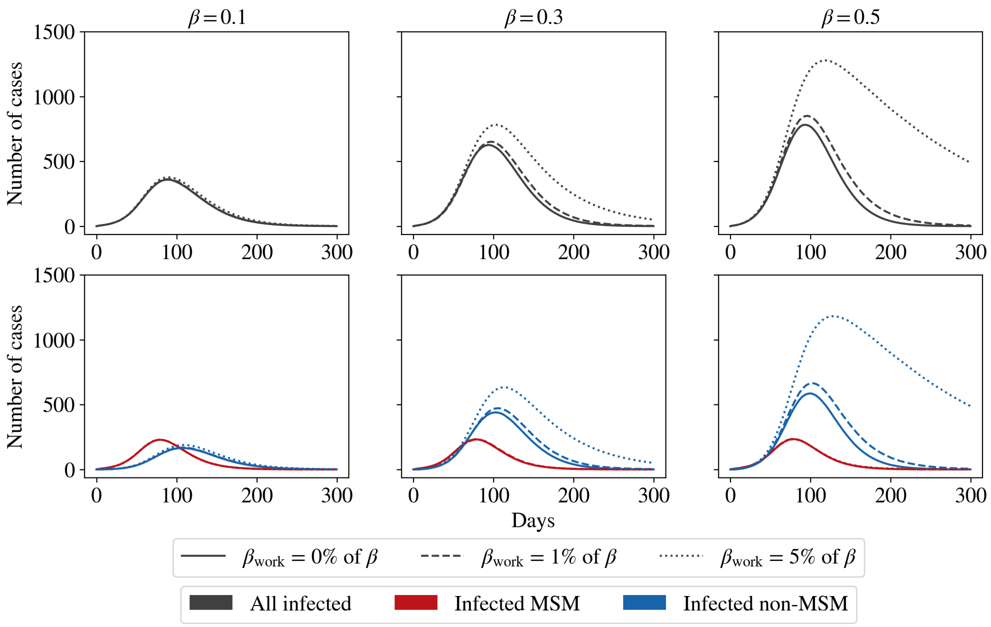


**Fig S1.3: Effects of weak workplace transmission on active infections in the extended population.** Just as in the original simulations, while a workplace contact multiplier of 1% does not affect the peak significantly, a multiplier of 5% leads to a much higher peak and a much heavier tail. As before, the curves are averages over 500 runs.

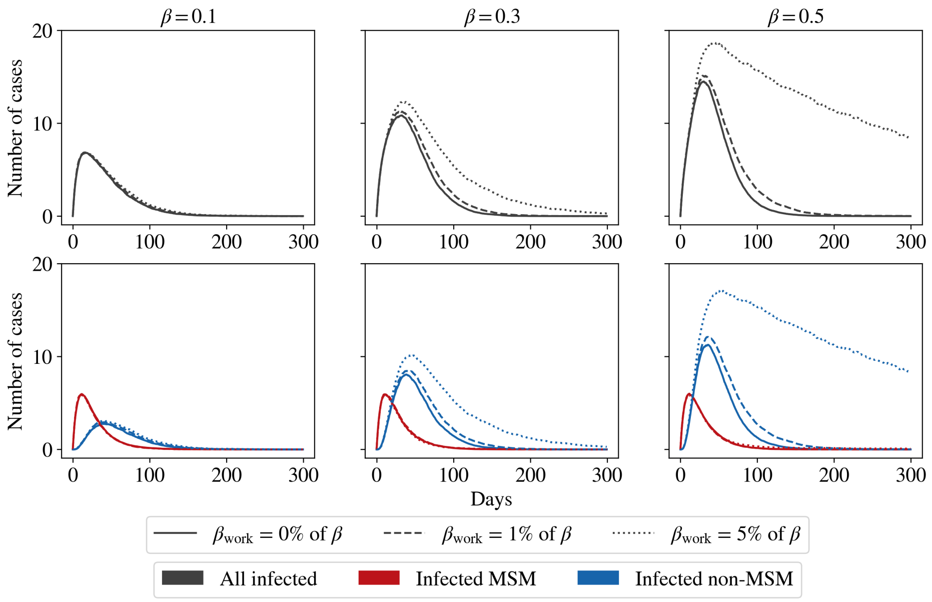


**Fig S1.4: Effects of weak workplace transmission on active infections in the extended population without an underlying MSM subnetwork.** We repeat the simulations of Fig. S1.3, but with $\mu = 0$, meaning that the disease does not spread in the MSM subnetwork. This result is identical to that from Fig. S7.1, as is expected. As before, the curves are averages over 500 runs.

### S2 Appendix: Effect of bisexual partnerships on disease spread

We now study the effect of introducing female sexual partners to a subset of the MSM agents in our population. Studies have shown that close to a third of the MSM population is bisexual [1, 2]. Of these, around 75% have a predominant female partner who is their wife. Consequently, we have also run simulations in which 33% of MSMs are bisexual.

We begin by constructing the population (with its associated MSM subnetwork) as described in Section 2.4 of the main paper. Once this is done, we assign 33% of the agents as bisexual, indicating that in addition to their MSM contacts drawn from the distribution as described in Section 2.4, a single female household member is further designated as their female sexual partner. We assume that these female partners do not participate in additional sexual partnerships beyond their associated bisexual MSM agent and therefore do not contribute to further sexual transmission. They can, however, transmit the disease non-sexually to other household contacts.

We emphasise that this representation is intentionally simplified; we do not account for heterogeneity in the number and type of female sexual partnerships and sexual-contact frequencies. For example, Ref. [2] has shown that despite the predominant sexual partner of bisexual MSMs being their wife, 13% of MSMs have also reported sex with women outside of marriage including with commercial sex workers. We nevertheless acknowledge that incorporating such features could alter the coupling between the MSM and non-MSM networks. However, we believe this effect to be subdominant.

Sexual encounters occur as described in the main paper, however bisexual MSM agents' contacts now include their female sexual partner. Given the paucity of evidence regarding the difference in transmission between anal and vaginal routes of sexual transmission, we have assumed that the probability of sexual transmission per encounter remains the same between the MSM and their female partner as between MSMs. However, this parameter can be varied as new evidence is obtained in the future.

Our results for this population, shown in Fig. S2.1, show that the introduction of bisexual partnerships in the MSM sub-network slows down the disease spread in the MSM subnetwork and consequently in the non-MSM network as well. We understand this as follows: the designation of a subset of MSMs as bisexual (with female sexual contacts) effectively reduces the probability that these MSMs can transmit the disease to their MSM partners. In our model, since the female contacts themselves do not transmit the disease sexually to anyone else, these partners effectively play the role of "dead-ends" in the MSM sexual network. Thus, using a similar argument as in Appendix S1 where we studied the extended MSM sexual network, the presence of these dead-ends slows down disease spread.

The effects of this reduction in the non-MSM network depend on the interplay of $\beta$ and $\mu$. Since the spread in the non-MSM network depends on the number of infected MSMs, this spread is correspondingly slowed down as well. In general, for almost all the parameters chosen in this work, the inclusion of bisexual MSMs leads to a corresponding reduction in the number of cases in the non-MSM network. An important exception occurs when $\beta = 0.1$ and $\mu = 1.0$. In this case, we see a modest rise in the number of cases in the non-MSM subnetwork. The explanation for this is that the inclusion of female household partners can increase the household attack rate. However, this only happens when the household transmission rate ($\beta$) is low, and the probability of sexual transmission per encounter ($\mu$) is high. Thus, for low $\beta$ and high $\mu$, despite the slower spread, the total *number* of non-MSM cases could increase.

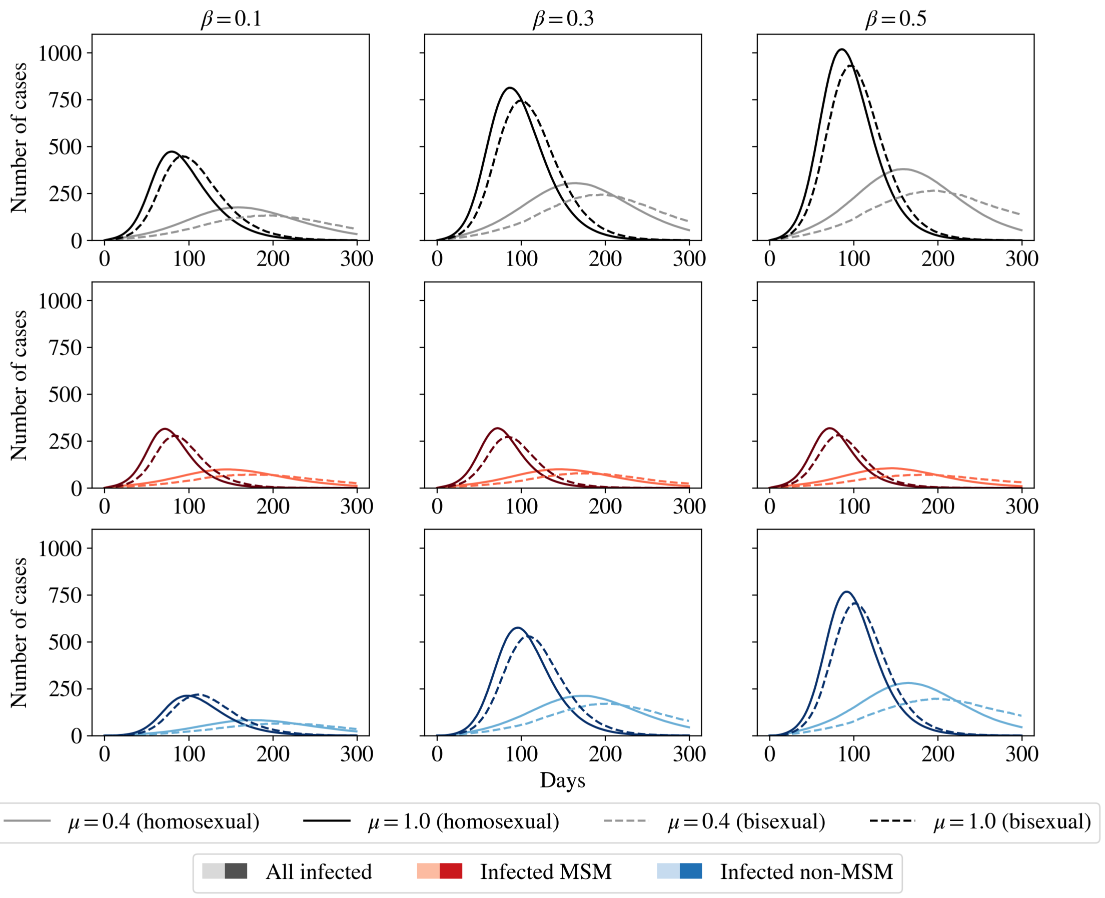


**Fig S2.1: Introducing bisexual MSMs to the population.** We re-run the same baseline runs from the main paper but with a population in which 33% of the MSMs have one household female sexual partner. For simplicity, we show only the results for $\mu = 0.4$ (dark lines) and $\mu = 1.0$ (faint lines) for the case in which sexual transmission amongst MSMs is exclusively homosexual (solid lines; same results as the main paper) and bisexual (dashed lines). We see that the introduction of bisexual contacts slows down the spread in the MSM-subnetwork and consequently in the non-MSM network. However, for low $\beta$ and high $\mu$ this could nevertheless lead to a marginally larger number of non-MSM cases, as it effectively increases the household attack rate in MSM households.

### S3 Appendix: Simulations for an initial infection seed of non-MSM agents

We consider the counterfactual scenario where a disease peak is observed due to an initial infection seed of 100 non-MSM agents. We run these simulations on a smaller population of 10,000 agents, of which 100 are MSMs. We find the results are qualitatively identical to the larger populations.

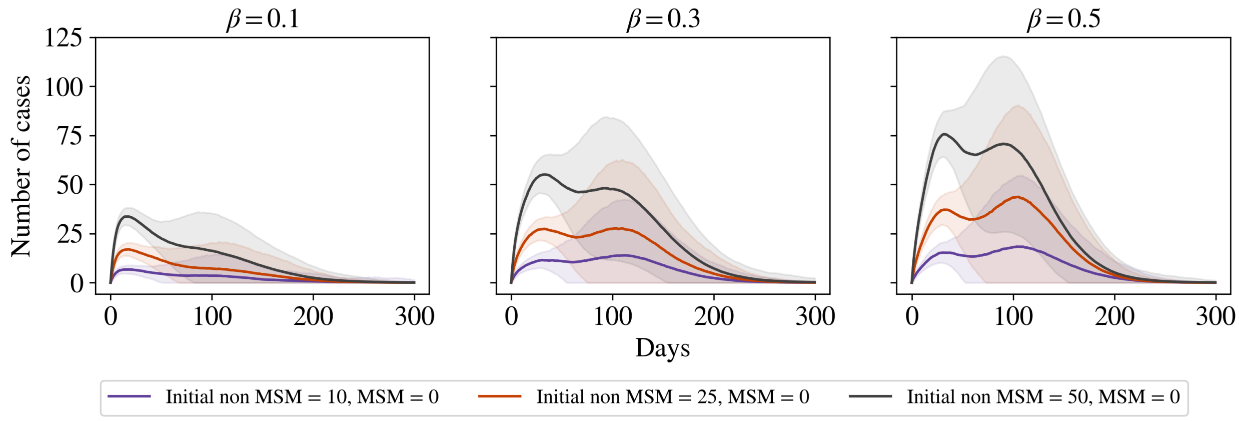


**Fig S3.1: Achieving similar disease peaks with an initial non-MSM seed.** We initialise infection with 10, 25, and 50 infected non-MSM agents. The broad, filled regions represent $1\sigma$ confidence intervals, indicating that entry into the MSM population is not guaranteed, especially at lower infection seeds. We stress that these results are dependent on the structure of the population. All results are averages over more than 500 stochastic runs.

We find that in order to achieve a peak in active infections that is comparable with the peak that would have occurred when the initial infection seed was 1% of MSMs, we would need to start with close to 50 non-MSM agents. These initial conditions lead to an interesting two-peak structure in the active infections. The first peak is due to infection in the non-MSM community. Given sufficient transmission, this can then cause the disease to enter the the MSM network, where it spreads much more rapidly, leading to the second, MSM-driven, peak. However, these results depend strongly on the structure of the population.

### S4 Appendix: Effect of sexual contact frequency on disease spread

In this section we consider the effect of varying the frequency at which MSMs engage in sexual activity within their sexual network. In the main paper we have considered that MSMs engage in a sexual encounter on average once a week, i.e. $\tau = 7$ days. (Since encounters are initiated independently per agent, this translates to each pair of MSMs who are sexual contacts of each other meeting twice a week on average.)

Here, we consider scenarios in which MSM agents engage in sexual activity (i) twice every week ($\tau = 3.5$ days, shown in Fig. S4.1), and (ii) once every 2 weeks ($\tau = 14$ days, shown in Fig. S4.2).

We find, as expected, that increasing the frequency of sexual encounters (i.e. reducing $\tau$) effectively increases the reproductive ratio for sexual contacts, as the rate of creating secondary infections via sexual transmission must be proportional to the frequency with which partners meet each other. We also find that in the range considered here, the increase in frequency can be offset by a corresponding *decrease* in the probability of sexual transmission (i.e., $\mu$) for the same reason.

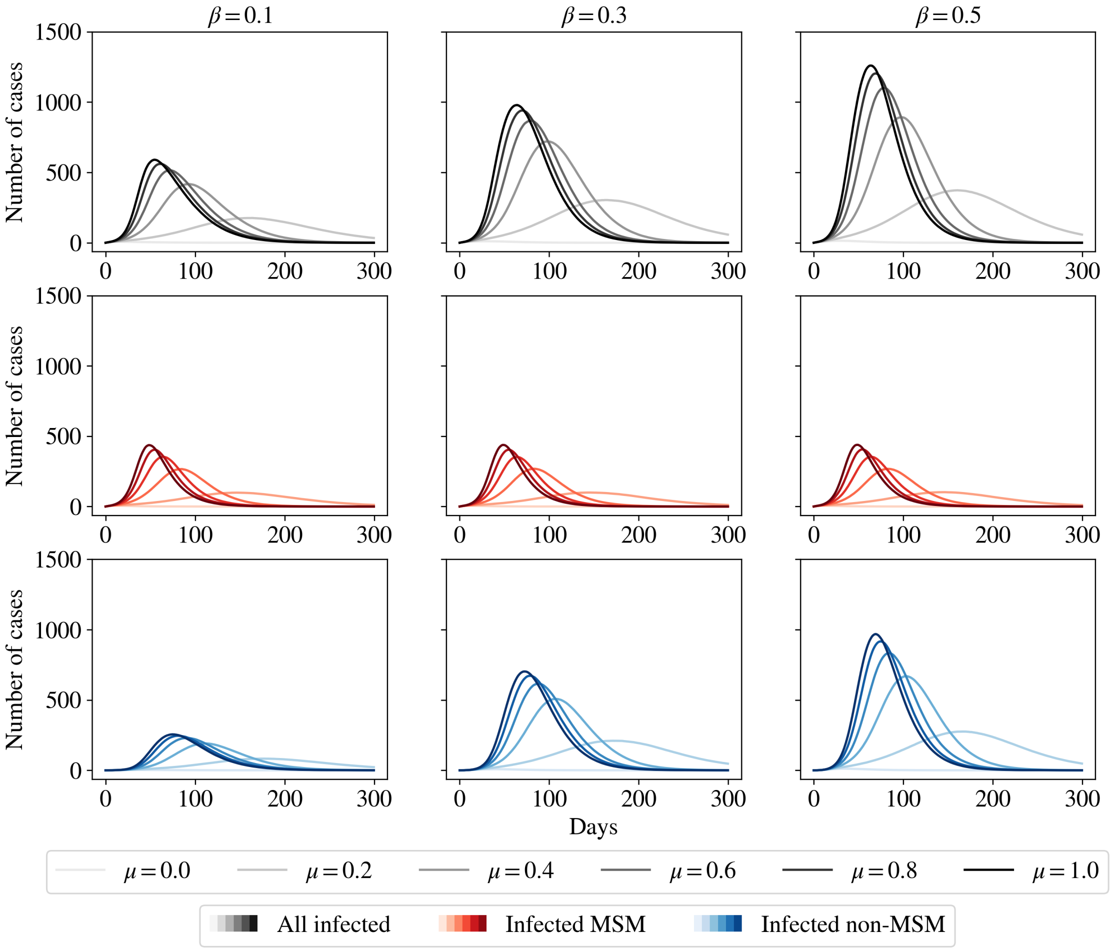


**Fig S4.1: Effects of increasing frequency of sexual contacts.** We re-run the same baseline runs from the main paper but increase the encounter frequency to twice a week. This leads to a much steeper rise in cases in the sexual network, which translates to an overall rise that is also steeper.

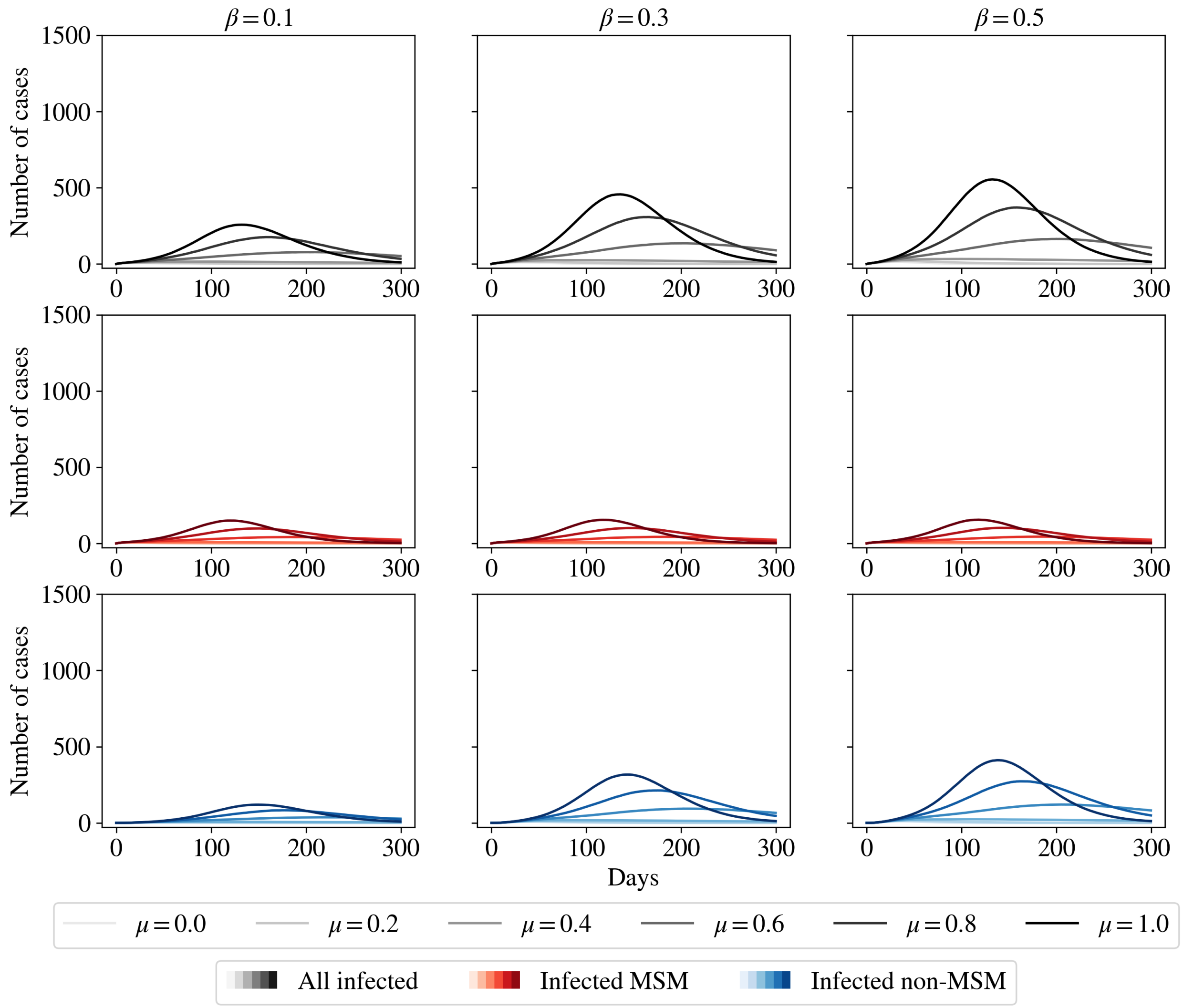


**Fig S4.2: Effects of decreasing the frequency of sexual contacts.** We re-run the same baseline runs from the main paper but *decrease* the meeting frequency to once in two weeks. This reduces the rise in cases in the sexual network, which consequently slows down the spread in the rest of the population.

### S5 Appendix: Additional results for vaccination strategies

In the main paper, we considered the case when 10 vaccines (VR=1%) were available per day, and were administered to MSMs in the population. Here, we consider the situation where there are 100 vaccines per day. At this rate, all MSM agents in our population would be vaccinated in a 10-day period. The vaccine is assumed to still be a single-dose vaccine, with an efficacy of 75% in reducing probability of contracting the disease.

We recreate Fig. 5 of the main paper using 100 vaccines per day (corresponding to a vaccination rate of 10% of the MSM population), as shown in Fig. S5.1. We see that all effects of different vaccination strategies are washed out at these high vaccination rates.

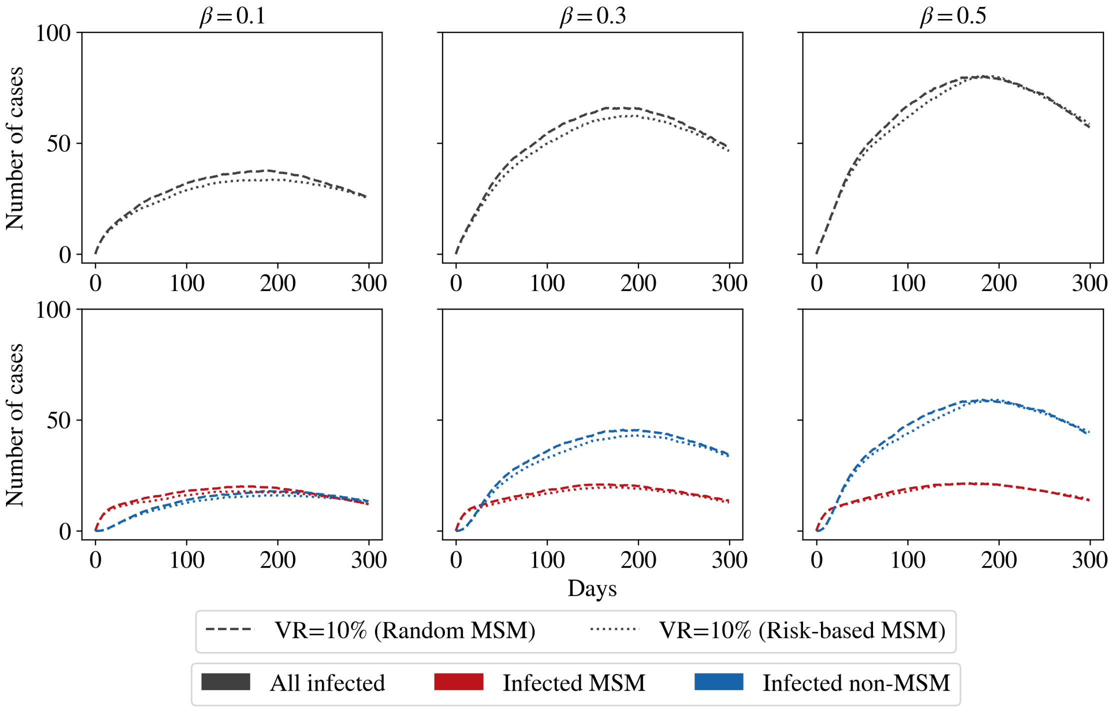


**Fig S5.1: Effects of vaccination strategies on disease peak.** Vaccinating 100 individuals per day is very efficient in reducing the peak of infection, with all differences in strategy being completely washed out. Again, the curves are averages over 500 runs.

We can now look at the combined effect of allowing for weak workplace transmission and a concurrent vaccination drive. In Fig. S5.2 we show results for different workplace transmission factors of 0%, 1%, and 5%, alongside a concurrent vaccination drive of 10 vaccines per day (VR=1% of MSMs). We see that vaccination greatly reduces the number of cases, reducing the peak from close to 1500 cases (see Fig. 7 of the main paper) to 750 cases in the worst case of a 5% workplace transmission. At higher vaccination rates of 100 vaccines per day (not shown here), the peak in active infections is effectively removed.

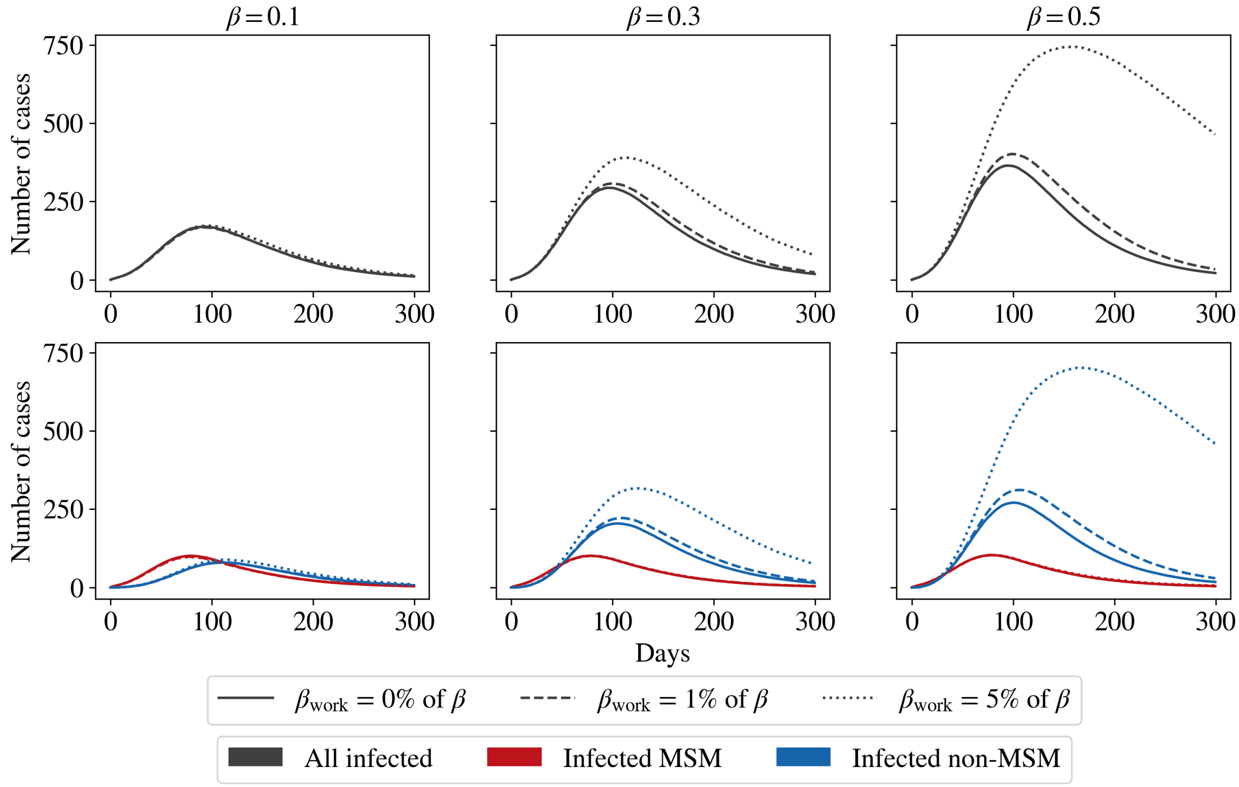


**Fig S5.2: Effects of a VR=1% and workplace transmission on disease peak.** A daily vaccination of 1% of the MSM population is effective at reducing the peak. However, at higher workplace transmission, a longer tail of infection persists. The curves are averages over 500 runs.

### S6 Appendix: Effect of varying $\tau_{SR}$ and $f_{trace}$ in ring vaccination

To study the effect of the self-reporting rates and contact tracing efficiencies on the overall reduction in cases because of a ring vaccination strategy, we run simulations for multiple parameters: first, we set $\tau_{SR} = 1$ day (corresponding to "immediate" reporting) and run simulations for all values of $f_{trace}$. Next, we set $f_{trace} = 80\%$ and show the results for all mean self-reporting delays. These plots can be seen in Figs. S6.1 and S6.2 respectively.

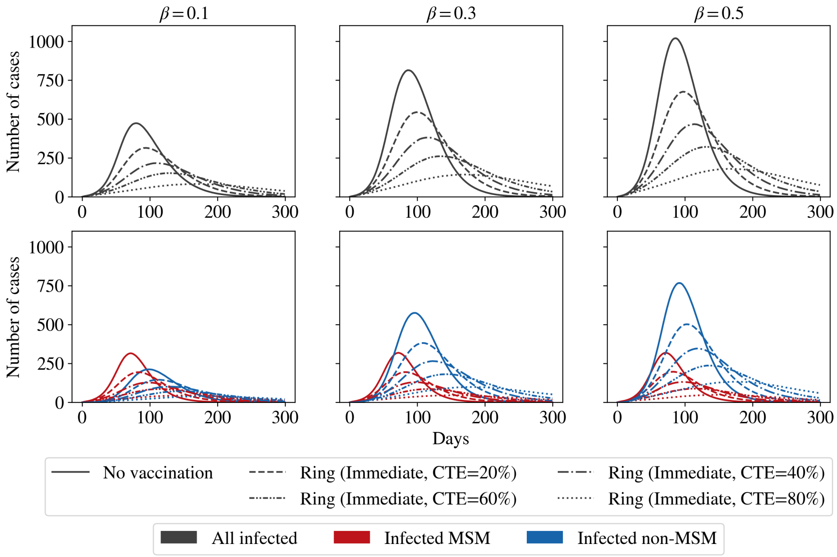


**Fig S6.1: Effect of contact tracing efficiency on ring vaccination.** Higher tracing efficiencies pull the peak down and shift it to later times. As before, the curves are averages over 500 runs.

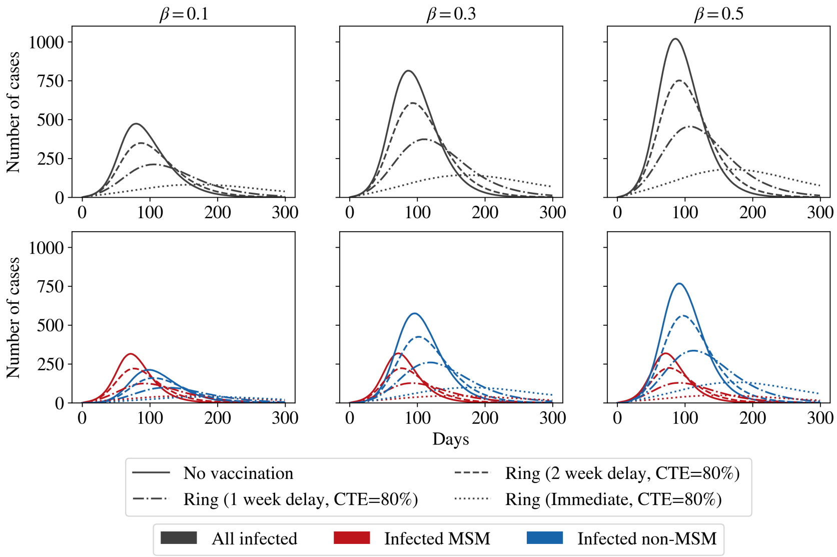


**Fig S6.2: Effect of self-reporting delay on ring vaccination.** The difference between immediate reporting and a 1-week delay is significant, but the effect is diminished when the delay is increased to 2 weeks. However, even at 2-week delays, 80% contact tracing efficiency can produce significant reductions in the active infections. As before, the curves are averages over 500 runs.

#### S7 Appendix: Primary and secondary transmission pathways

We first study the role that transmission in the MSM sexual network has on the spread of the disease. We do this by examining the same results as in Fig. 7 of the main paper, but with $\mu = 0$, effectively removing sexual contact as a possible channel for disease transmission. We see in Fig. S7.1 that in all cases the disease dies out without spreading. Thus, the much smaller MSM subnetwork is essential as it, along with the long generation time, contributes to sustaining the disease in the population.

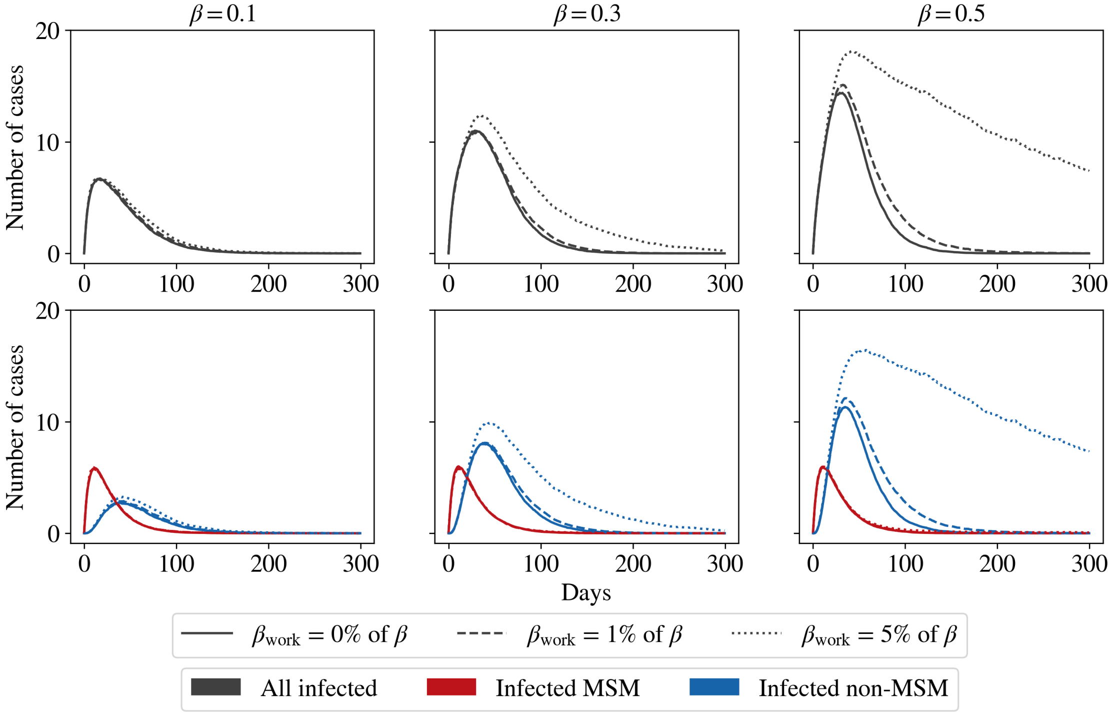


**Fig S7.1: Effects of weak workplace transmission on active infections without an underlying MSM subnetwork.** We repeat the simulations of Fig. 7 in the main paper, but with $\mu = 0$, meaning that the disease does not spread in the MSM subnetwork. We find that in every case, the disease dies out without spreading to the general population. As before, the curves are averages over 500 runs.

Additionally, to identify the source of the long tail in infections, we further show the results of Fig. 7 of the main paper, but divide the active infections into three classes: infections caused by MSMs (either through sexual transmission with their sexual contacts, or through non-sexual household transmission), infections caused by household members of MSMs, and infections caused by secondary contacts whom we identify as workplace contacts of MSMs and their household members. In Fig. S7.2 we compare the cases associated with these three classes of "infectors" on the population. We see from this graph that the long tail arises almost exclusively from the secondary contacts of MSMs and their households. Allowing for a small possibility of workplace transmission thus allows the disease to escape the initially high-risk transmission network and persist through broader workplace-mediated transmission chains in the general population, which sustain long epidemic tails even after direct transmission within the MSM sub-network has declined.

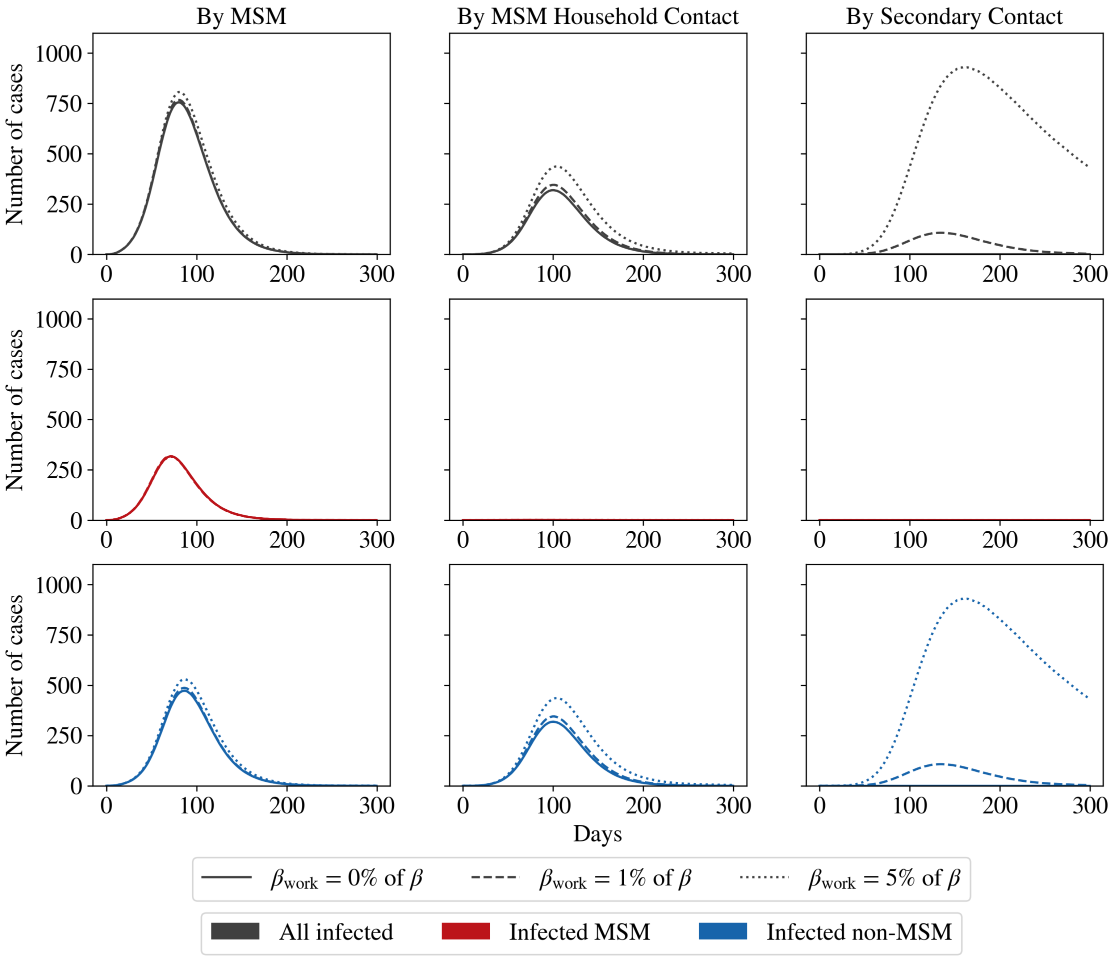


**Fig S7.2: Cases categorised by infector type.** We repeat the simulations of Fig. 7 in the main paper, but categorise the active infections based on the type of infecting agent. We divide the population into three categories: MSMs, their household contacts, and all workplace contacts of MSMs and their households. These are represented by the three columns. For each category, we identify the cases in the total population (top row), MSM population (middle row), and non-MSM population (bottom row). We find that the long tail in infections is driven by workplace contacts of MSMs and their households, which allows the disease to persist even after it has died out in the high-risk MSM sub-network. As before, the curves are averages over 500 runs.
